# Durability of protection from original monovalent and bivalent COVID-19 vaccines against COVID-19-associated hospitalization and severe in-hospital outcomes among adults in the United States — September 2022–August 2023

**DOI:** 10.1101/2024.01.07.24300910

**Authors:** Jennifer DeCuir, Diya Surie, Yuwei Zhu, Adam S. Lauring, Manjusha Gaglani, Tresa McNeal, Shekhar Ghamande, Ithan D. Peltan, Samuel M. Brown, Adit A. Ginde, Aimee Steinwand, Nicholas M. Mohr, Kevin W. Gibbs, David N. Hager, Harith Ali, Anne Frosch, Michelle N. Gong, Amira Mohamed, Nicholas J. Johnson, Vasisht Srinivasan, Jay S. Steingrub, Akram Khan, Laurence W. Busse, Abhijit Duggal, Jennifer G. Wilson, Nida Qadir, Steven Y. Chang, Christopher Mallow, Jennie H. Kwon, Matthew C. Exline, Nathan I. Shapiro, Cristie Columbus, Ivana A. Vaughn, Mayur Ramesh, Basmah Safdar, Jarrod M. Mosier, Jonathan D. Casey, H. Keipp Talbot, Todd W. Rice, Natasha Halasa, James D. Chappell, Carlos G. Grijalva, Adrienne Baughman, Kelsey N. Womack, Jillian P. Rhoads, Sydney A. Swan, Cassandra Johnson, Nathaniel Lewis, Sascha Ellington, Meredith McMorrow, Wesley H. Self, Investigating Respiratory Viruses in the Acutely Ill (IVY) Network

**Affiliations:** National Center for Immunization and Respiratory Diseases, Centers for Disease Control and Prevention (CDC), Atlanta, Georgia; Department of Biostatistics, Vanderbilt University Medical Center, Nashville, Tennessee; Departments of Internal Medicine and Microbiology and Immunology, University of Michigan, Ann Arbor, Michigan; Baylor Scott and White Health, Temple and Dallas, Texas, and Texas A&M University College of Medicine, Temple, Texas; Baylor Scott and White Health, and Baylor College of Medicine, Temple, Texas; Baylor Scott and White Health, Texas A&M University College of Medicine, Temple, Texas; Department of Medicine, Intermountain Medical Center, Murray, Utah and University of Utah, Salt Lake City, Utah; Department of Emergency Medicine, University of Colorado School of Medicine, Aurora, Colorado; Department of Emergency Medicine, University of Colorado School of Medicine, Aurora, CO; University of Iowa, Iowa City, Iowa; Department of Medicine, Wake Forest School of Medicine, Winston-Salem, North Carolina; Department of Medicine, Johns Hopkins University School of Medicine, Baltimore, Maryland; Department of Emergency Medicine, Hennepin Healthcare Research Institute, Hennepin Healthcare System, Minneapolis, Minnesota; Department of Medicine, Montefiore Medical Center, Albert Einstein College of Medicine, Bronx, New York; Department of Emergency Medicine and Division of Pulmonary, Critical Care and Sleep Medicine, University of Washington, Seattle, Washington; Department of Emergency Medicine University of Washington, Seattle, Washington; Department of Medicine, Baystate Medical Center, Springfield, Massachusetts; Department of Medicine, Oregon Health and Sciences University, Portland, Oregon; Department of Medicine, Emory University, Atlanta, Georgia; Department of Medicine, Cleveland Clinic, Cleveland, Ohio; Department of Emergency Medicine, Stanford University School of Medicine, Stanford, California; Department of Medicine, University of California-Los Angeles, Los Angeles, California; Department of Medicine, University of Miami, Miami, Florida; Department of Medicine, Washington University, St. Louis, Missouri; Department of Medicine, The Ohio State University, Columbus, Ohio; Department of Emergency Medicine, Beth Israel Deaconess Medical Center, Boston, Massachusetts; Baylor Scott & White Health, Dallas, Texas; Department of Public Health Sciences, Henry Ford Health, Detroit, Michigan; Division of Infectious Diseases, Henry Ford Health, Detroit, Michigan; Department of Emergency Medicine, Yale University School of Medicine, New Haven, Connecticut; Department of Emergency Medicine, University of Arizona, Tucson, Arizona; Department of Medicine, Vanderbilt University Medical Center, Nashville, Tennessee; Departments of Medicine and Health Policy, Vanderbilt University Medical Center, Nashville, Tennessee; Department of Pediatrics, Vanderbilt University Medical Center, Nashville, Tennessee; Department of Health Policy, Vanderbilt University Medical Center, Nashville, Tennessee; Department of Emergency Medicine, Vanderbilt University Medical Center, Nashville, Tennessee; Vanderbilt Institute for Clinical and Translational Research, Vanderbilt University Medical Center, Nashville, Tennessee; Vanderbilt Institute for Clinical and Translational Research, and Department of Emergency Medicine, Vanderbilt University Medical Center, Nashville, Tennessee

## Abstract

**Objective:** To evaluate the durability of protection provided by original monovalent and bivalent COVID-19 vaccination against COVID-19-associated hospitalization and severe in-hospital outcomes.

**Design:** Multicenter case-control design with prospective enrollment

**Setting:** 26 hospitals in 20 US states

**Participants:** Adults aged ≥18 years admitted to hospital with COVID-19-like illness from 8 September 2022 to 31 August 2023

**Main outcome measures:** The main outcomes were absolute and relative vaccine effectiveness of original monovalent and bivalent COVID-19 vaccines against COVID-19-associated hospitalization and severe in-hospital outcomes, including advanced respiratory support (defined as receipt of high-flow nasal cannula, non-invasive ventilation, or invasive mechanical ventilation [IMV]) and IMV or death. Vaccine effectiveness was estimated using multivariable logistic regression, in which the odds of vaccination (versus being unvaccinated or receiving original monovalent vaccination only) were compared between COVID-19 case patients and control-patients. Bivalent vaccine effectiveness analyses were stratified by time since dose receipt.

**Results:** Among 7028 adults without immunocompromising conditions, 2924 (41.6%) were COVID-19 case patients and 4104 (58.4%) were control patients. Compared to unvaccinated patients, absolute vaccine effectiveness against COVID-19-associated hospitalization was 6% (-7% to 17%) for original monovalent doses only (median time since last dose [IQR] = 421 days [304–571]), 52% (39% to 61%) for a bivalent dose received 7–89 days earlier, and 13% (-10% to 31%) for a bivalent dose received 90–179 days earlier. Absolute vaccine effectiveness against COVID-19-associated advanced respiratory support was 31% (15% to 45%) for original monovalent doses only, 66% (47% to 78%) for a bivalent dose received 7–89 days earlier, and 33% (-1% to 55%) for a bivalent dose received 90–179 days earlier. Absolute vaccine effectiveness against COVID-19-associated IMV or death was 51% (34% to 63%) for original monovalent doses only, 61% (35% to 77%) for a bivalent dose received 7–89 days earlier, and 50% (11% to 71%) for a bivalent dose received 90–179 days earlier.

**Conclusion:** When compared to original monovalent vaccination only, bivalent COVID-19 vaccination provided additional protection against COVID-19-associated hospitalization and certain severe in-hospital outcomes within 3 months of dose receipt. By 3-6 months, protection from a bivalent dose declined to a level similar to that remaining from original monovalent vaccination only. Although no protection remained from original monovalent vaccination against COVID-19-associated hospitalization, it provided durable protection against severe in-hospital outcomes >1 year after receipt of the last dose, particularly against IMV or death.

**SUMMARY BOX:** *What is already known on this topic:*

- On September 1, 2022, bivalent mRNA COVID-19 vaccination was recommended for US adults who had completed at least an original monovalent COVID-19 primary series.
- Early estimates of bivalent vaccine effectiveness are available for the period soon after dose receipt; however fewer data exist on their durability of protection and effectiveness against severe outcomes.

*What this study adds:*

- When compared to original monovalent vaccination only, bivalent mRNA COVID-19 vaccination provided additional protection against COVID-19-associated hospitalization and certain severe in-hospital outcomes within 3 months of dose receipt. By 3-6 months, protection from a bivalent dose declined to a level similar to that remaining from original monovalent vaccination only.
- Although no protection remained from original monovalent vaccination against COVID-19-associated hospitalization, it provided durable protection against severe in-hospital outcomes >1 year after receipt of the last dose, particularly against invasive mechanical ventilation or death.

## INTRODUCTION

In December 2020, monovalent COVID-19 vaccines designed against the ancestral strain of SARS-CoV-2 (original monovalent vaccines) were introduced in the United States to prevent COVID-19-associated morbidity and mortality. While these vaccines successfully prevented COVID-19-associated hospitalization and death [1,2], their effectiveness declined over time due to several factors, including waning vaccine-induced immunity and the emergence of the immune evasive Omicron variant [3,4]. To address these factors, on September 1, 2022, the Advisory Committee on Immunization Practices at the US Centers for Disease Control and Prevention (CDC) recommended a bivalent mRNA COVID-19 dose, designed to protect against both the ancestral strain and Omicron BA.4/5 lineages, for persons who had completed at least an original monovalent COVID-19 primary series [5]. With implementation of this recommendation, original monovalent mRNA COVID-19 vaccines were no longer recommended for use as booster doses in the US [6]. In April 2023, COVID-19 vaccine recommendations were updated to state that bivalent vaccines should be used for all mRNA COVID-19 vaccine doses [7].

Early estimates showed that bivalent mRNA COVID-19 vaccines were effective against COVID-19-associated hospitalization shortly after administration [8–11]. However, fewer data exist on their durability of protection and their effectiveness against severe outcomes, particularly in the setting of residual protection remaining from original monovalent vaccination. Numerous Omicron lineages have also emerged in the US since the bivalent vaccine rollout, and the impact of these lineages on vaccine effectiveness is unclear. While neutralization studies suggest that the XBB.1.5 lineage is among the most immune evasive to date, few studies have measured bivalent vaccine effectiveness during the period of XBB.1.5 predominance [12–15].

To address these gaps, we evaluated bivalent mRNA COVID-19 vaccine effectiveness against COVID-19-associated hospitalization and severe in-hospital outcomes up to 6 months after dose receipt. Results were stratified by time from bivalent dose receipt to illness onset and Omicron lineage period to better differentiate the impact of waning vaccine-induced immunity and viral immune evasion on bivalent vaccine effectiveness.

## METHODS

### Setting and design

This case-control analysis was conducted by the Investigating Respiratory Viruses in the Acutely Ill (IVY) Network, comprised of 26 hospitals in 20 US states in collaboration with the CDC. The current analysis included adults admitted to IVY Network hospitals from 8 September 2022 to 31 August 2023. Vaccine effectiveness for the prevention of COVID-19-associated hospitalization and severe in-hospital outcomes was estimated for the original monovalent and bivalent COVID-19 vaccines authorized for use in the US during the analysis period. These activities were determined to be public health surveillance with waiver of informed consent by institutional review boards at CDC and each enrolling site, and were conducted in accordance with applicable federal law and CDC policy (45 C.F.R. part 46.102(l)(2), 21 C.F.R. part 56; 42 U.S.C. §241(d); 5 U.S.C. §552a; 44 U.S.C. §3501 et seq).

### Participants

Site personnel prospectively enrolled patients aged ≥18 years admitted to IVY Network hospitals who met a COVID-19-like illness case definition and received SARS-CoV-2 clinical testing. COVID-19-like illness was defined as ≥1 of the following signs and symptoms: fever, cough, shortness of breath, new or worsening findings on chest imaging consistent with pneumonia, and hypoxemia (defined as an oxygen saturation [SpO_2_] <92% or supplemental oxygen use for patients without chronic oxygen needs, or escalation of oxygen therapy for patients on chronic supplemental oxygen). Case patients tested positive for SARS-CoV-2 by nucleic acid amplification test or antigen test within 10 days of illness onset and within 3 days of hospital admission. Control patients tested negative for SARS-CoV-2 by real-time reverse transcription polymerase chain reaction (RT-PCR) within 10 days of illness onset and within 3 days of hospital admission. Case-control status was determined by results of both clinical SARS-CoV-2 testing at the admitting hospital, as well as standardized, central laboratory testing for SARS-CoV-2 by RT-PCR. Patients who tested positive for SARS-CoV-2 by either clinical testing or central laboratory testing were classified as cases, while patients who tested negative by both clinical and central laboratory testing were classified as controls. Case patients who tested positive for influenza or RSV were excluded from analyses, while control patients who tested positive for influenza were also excluded due to potential correlation between COVID-19 and influenza vaccination behaviors [16]. Case and control patients were enrolled within two weeks of one another at each site in approximately a 1:1 ratio and were not matched on other patient-level characteristics.

### Data collection

Trained personnel at enrolling sites collected data on patient demographics, chronic medical conditions, COVID-19 vaccination status, and severe in-hospital outcomes through patient or proxy interview and medical chart abstraction. Chronic medical conditions were grouped into nine categories: cardiovascular, pulmonary, renal, endocrine, gastrointestinal, hematologic, neurologic, autoimmune, and immunocompromising (see supplemental table S1). Verification of COVID-19 vaccination status was performed using hospital electronic medical records, state vaccine registries, and vaccination cards (when available). Patients were followed from admission until discharge, death, or hospital day 28 (whichever occurred first).

### Classification of vaccination status

To assess bivalent vaccine effectiveness, patients were classified into four vaccination status groups: 1) unvaccinated (no COVID-19 vaccine doses received), 2) vaccinated with original monovalent doses only (receipt of any combination of 1–4 doses of original monovalent vaccine: mRNA-1273 [Moderna], BNT162b2 [Pfizer-BioNTech], Ad26.COV2.S [Janssen], NVX-CoV2373 [Novavax]), 3) vaccinated with one bivalent mRNA dose 7–89 days before illness onset, and 4) vaccinated with one bivalent mRNA dose 90–179 days before illness onset. Patients were classified as bivalent vaccinated if they received one dose of either mRNA-1273.222 (Moderna) or BNT1262b2 bivalent (Pfizer-BioNTech), regardless of the number of previous original monovalent doses received. Patients were excluded if they received >1 bivalent dose, if they received any COVID-19 vaccine dose <7 days before illness onset, or if they received only one dose of original monovalent Moderna, Pfizer, or Novavax vaccine. Estimates of bivalent vaccine effectiveness ≥180 days after dose receipt lacked sufficient precision for reporting; therefore, patients who received a bivalent dose ≥180 days before illness onset were also excluded. The main analysis was restricted to patients without immunocompromising conditions (described in supplementary appendix B). A sub-analysis of bivalent vaccine effectiveness among patients with immunocompromising conditions is available in the supplementary materials (supplemental table 1, supplemental figure 5).

### Clinical outcomes

The primary outcome in this analysis was COVID-19-associated hospitalization, defined as hospital admission with COVID-19-like illness and laboratory-confirmed SARS-CoV-2 infection as described above. Secondary outcomes were also included to assess vaccine effectiveness against severe COVID-19-associated disease through hospital day 28. These outcomes were: 1) supplemental oxygen therapy, defined as any receipt of supplemental oxygen for patients with no chronic oxygen use or escalation of oxygen use for patients on chronic supplemental oxygen; 2) advanced respiratory support, defined as new receipt of high-flow nasal cannula (HFNC), non-invasive ventilation (NIV), or invasive mechanical ventilation (IMV); 3) acute organ failure, defined as either respiratory failure (new receipt of HFNC, NIV, or IMV), cardiovascular failure (receipt of vasopressors), or renal failure (new receipt of renal replacement therapy); 4) intensive care unit (ICU) admission; and 5) IMV or death. Patients on home IMV were not eligible for the supplemental oxygen therapy or advanced respiratory support outcomes (outcomes are fully described in supplementary appendix B).

### Molecular diagnosis and sequencing

Nasal swabs were obtained from enrolled patients and tested for SARS-CoV-2, influenza, and RSV by RT-PCR at a central laboratory at Vanderbilt University Medical Center (Nashville, Tennessee). All specimens that tested positive for SARS-CoV-2 by central RT-PCR testing were submitted to the University of Michigan (Ann Arbor, Michigan) for viral whole genome sequencing. Sequencing was performed using the ARTIC Network protocol on either an Oxford Nanopore Technologies GridION or Illumina Nextseq 1000 instrument. SARS-CoV-2 lineages were reported using the PANGO (phylogenetic assignment of named global outbreak lineages) nomenclature [17]. Detailed laboratory methods are described in Supplementary Appendix B. SARS-CoV-2 lineages were considered predominant if they were detected in ≥50% of sequenced specimens during a given admission week.

### Statistical analysis

Bivalent and original monovalent vaccine effectiveness against COVID-19-associated hospitalization was calculated using multivariable logistic regression, in which the odds of COVID-19 vaccination were compared between COVID-19 case patients and control patients. Estimates of vaccine effectiveness were generated using two different comparator groups. First, absolute vaccine effectiveness was calculated for both bivalent and original monovalent vaccines using unvaccinated patients as the reference group. Second, relative vaccine effectiveness was calculated for bivalent vaccines using patients who received original monovalent vaccination only as the reference group. Logistic regression models were adjusted for age (18-49, 50-64, ≥65), sex (male, female), self-reported race and Hispanic ethnicity (non-Hispanic white, non-Hispanic Black, Hispanic or Latino, non-Hispanic other race, other), admission date in biweekly intervals, and U.S. Department of Health and Human Services region of the admitting hospital. Post hoc, we evaluated other variables as potential covariates, including number of chronic medical condition categories and known previous Omicron infection, defined as either self-reported or documented SARS-CoV-2 infection on or after 26 December 2021. None of these other variables resulted in an absolute change in the adjusted odds ratio of >5% when added to the prespecified model and were not included in the final models (supplemental table 2) [18]. Vaccine effectiveness was calculated as (1 − adjusted odds ratio) x 100%.

Overall estimates of bivalent and original monovalent vaccine effectiveness against COVID-19-associated hospitalization were stratified by age (18–64, ≥65 years), number of chronic medical condition categories (<3, ≥3), bivalent vaccine product type (Moderna, Pfizer), and Omicron lineage period (BA.4/5, XBB.1.5). Cut-off dates for each Omicron lineage period were determined using IVY Network sequencing data, with BA.4/5 being predominant from 8 September 2022 to 26 November 2022 and XBB.1.5 from 22 January 2023 to 27 May 2023. Analyses assessing bivalent and original monovalent vaccine effectiveness against COVID-19-associated in-hospital outcomes were conducted using the methods described for COVID-19-associated hospitalization, with cases limited to those who met each severe in-hospital outcome definition. Analyses were conducted using SAS (version 9.4; SAS Institute).

### Patient and public involvement

Patients were not involved in the original design of this study. Throughout the conduct of the study, patients were routinely engaged in the work via structured interviews with site personnel, which included questions about COVID-19 and COVID-19 vaccines.

## RESULTS

### Description of participants in vaccine effectiveness analyses

Between 8 September 2022 and 31 August 2023, 11797 patients admitted with COVID-19-like illness were enrolled in the IVY Network from 26 hospitals in 20 US states (supplemental figure S1). A total of 2374 (20.1%) patients met exclusion criteria and were removed from the analysis. Among the remaining 9423 patients, there were 7028 without immunocompromising conditions (included in the main analysis) and 2395 with immunocompromising conditions (sub-analysis available in the supplementary materials).

Among the patients without immunocompromising conditions, 2924 (41.6%) were COVID-19 case patients and 4104 (58.4%) were control patients (table 1). The median age was 67 years (interquartile range [IQR] 56–78), 3573 (50.8%) patients were female, 1606 (22.9%) were non-Hispanic Black, and 802 (11.4%) were Hispanic. A total of 1545 (22.0%) patients were unvaccinated, 4191 (59.6%) were vaccinated with original monovalent doses only, 647 (9.2%) were vaccinated with one bivalent dose 7–89 days before illness onset, and 645 (9.2%) were vaccinated with one bivalent dose 90–179 days before illness onset. Compared with patients who received either original monovalent or bivalent vaccine doses, unvaccinated patients were younger (p<0.0001) and more likely to be non-Hispanic Black (p=0.0338) or Hispanic (p=0.0002).

**Table 1.**
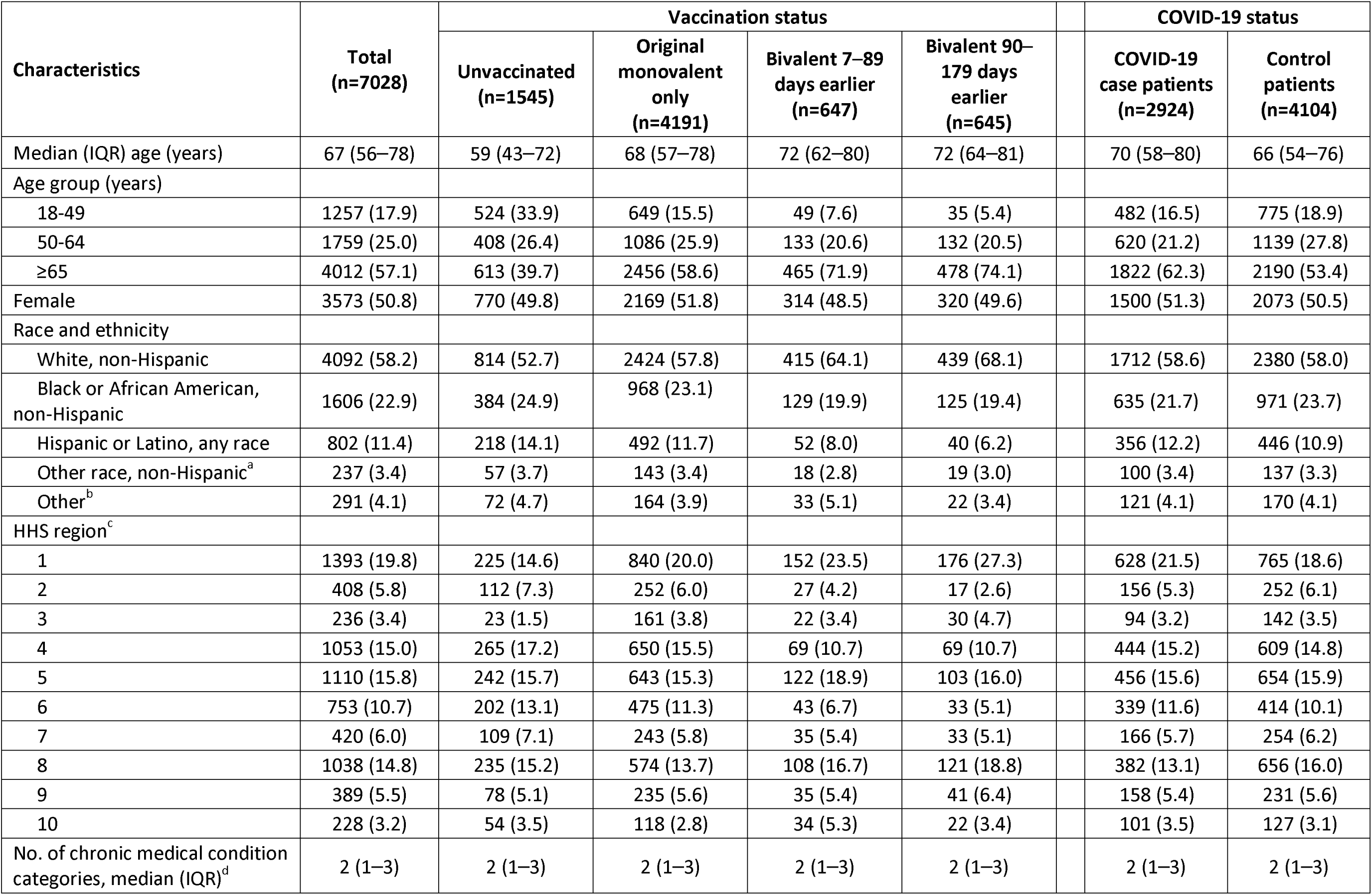
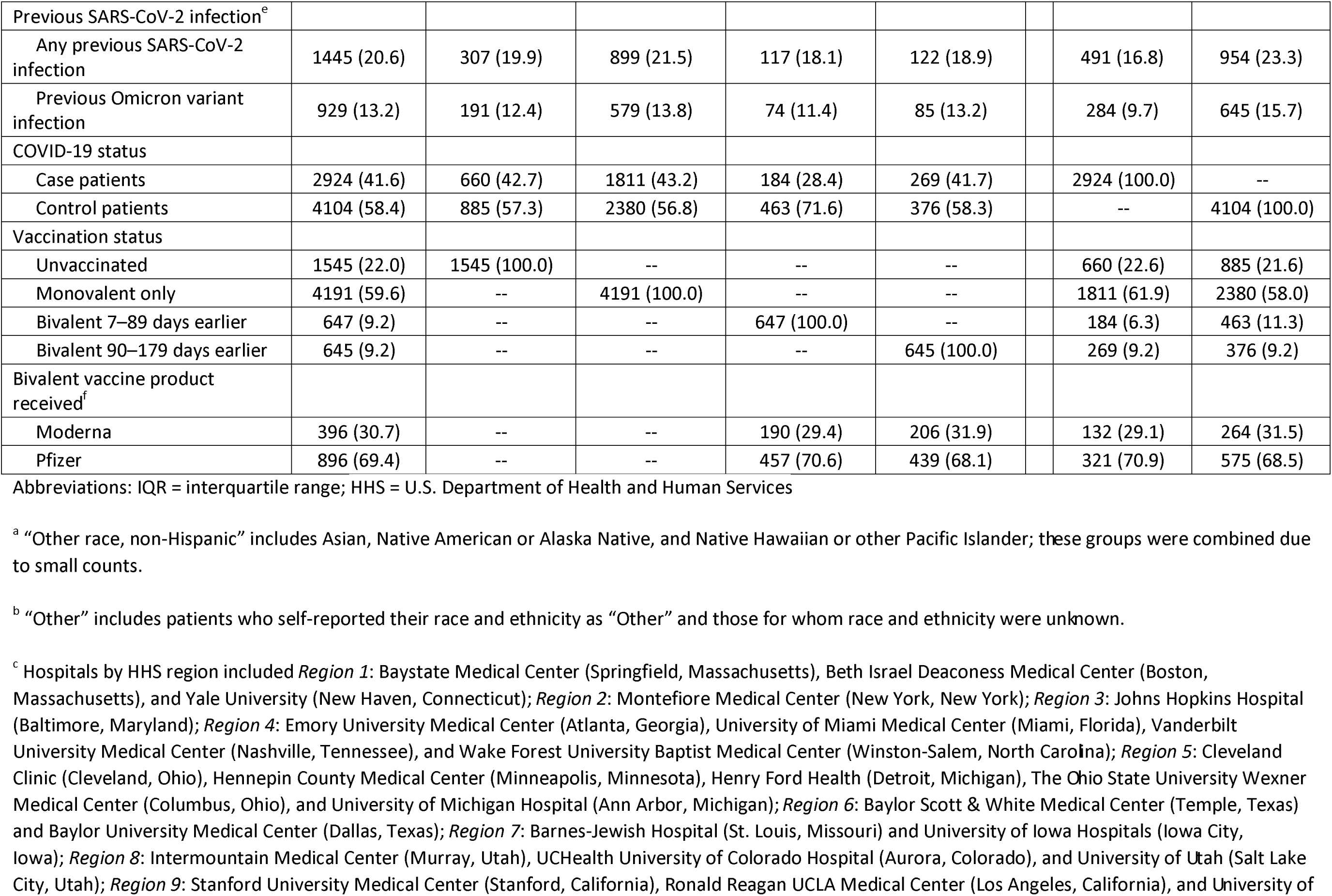
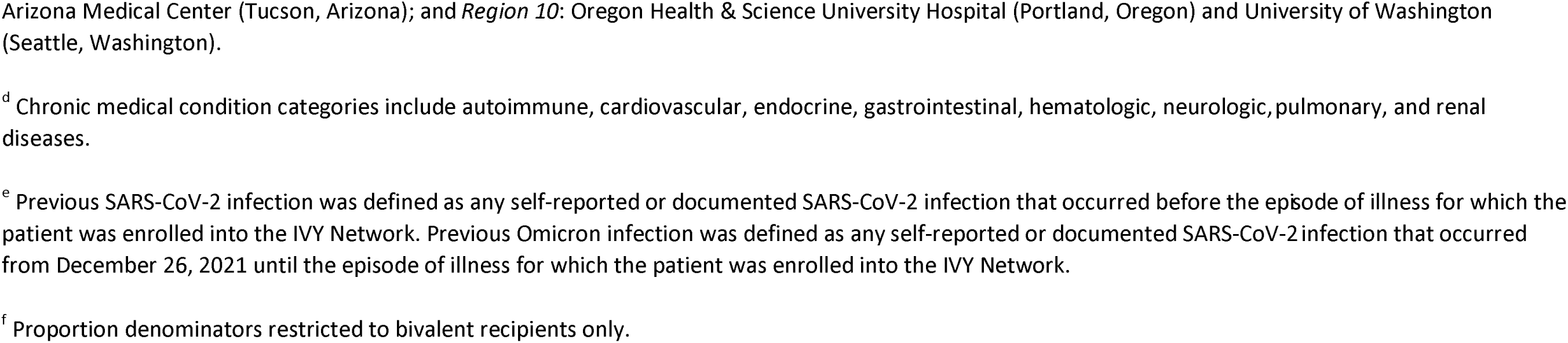
Characteristics of adults without immunocompromising conditions by vaccination and COVID-19 status admitted to one of 26 hospitals in 20 US states during 8 September 2022–31 August 2023. Values are numbers (column percentages) unless stated otherwise.

Viral whole genome sequencing was completed for 1779 of the 2924 case patients (60.8%), and a SARS-CoV-2 lineage was successfully identified for 1646 (56.3%), all of whom were infected with an Omicron lineage. Overall, 33 (2.0%) patients were infected with BA.2, 358 (21.7%) with BA.4/5, 346 (21.0%) with BQ.1, 701 (42.6%) with XBB.1.5, and 208 (12.6%) with other XBB lineages (figure 1). BA.4/5 was the predominant lineage from 8 September 2022 until 26 November 2022, and XBB.1.5 was predominant from 22 January 2023 to 27 May 2023.

**Figure 1.**
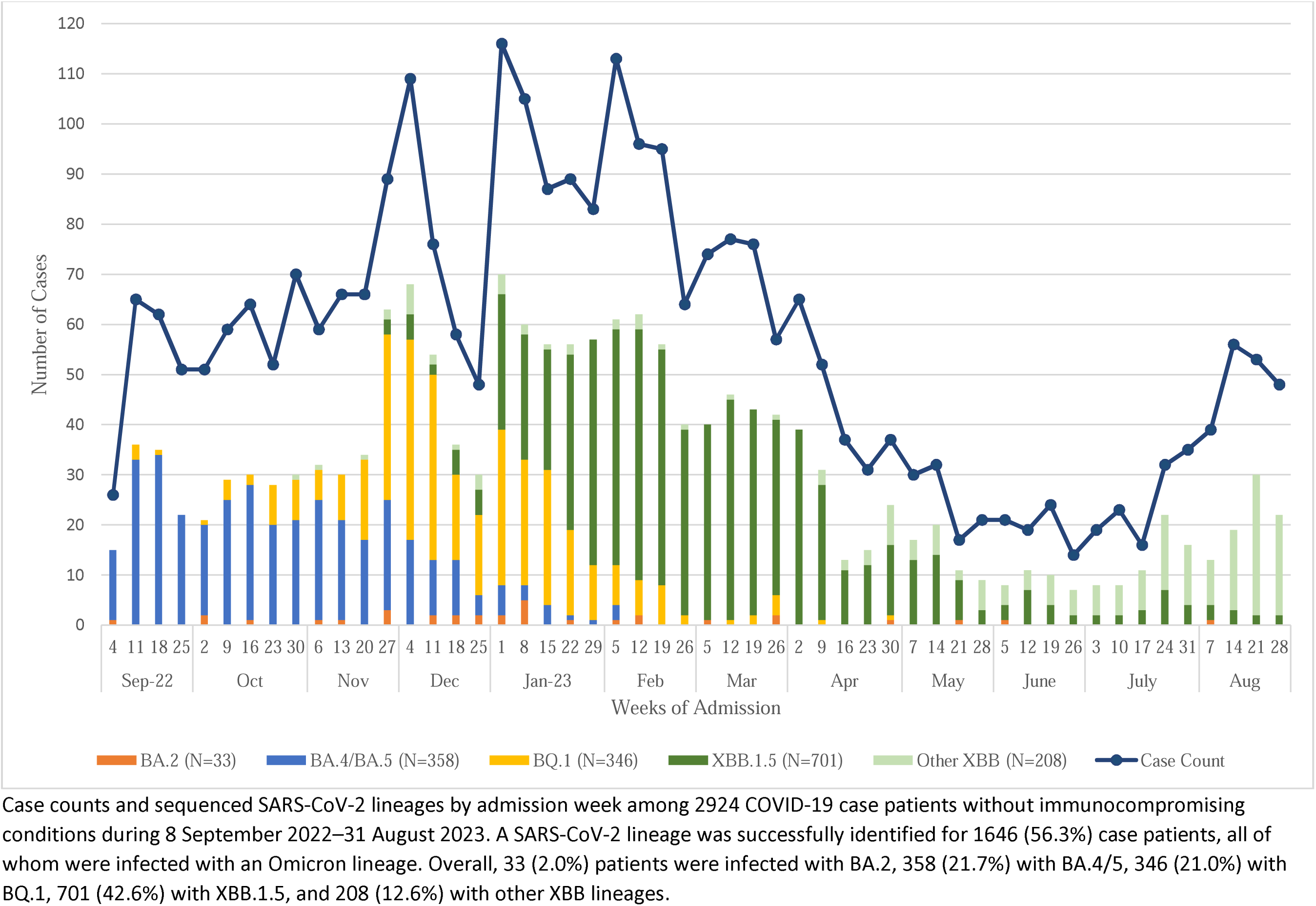

### Vaccine effectiveness against COVID-19-associated hospitalization

Among patients without immunocompromising conditions, the absolute effectiveness of original monovalent vaccination only against COVID-19-associated hospitalization was 6% (95% CI -7% to 17%), with a median time since last dose of 421 days (IQR 304–571). Bivalent vaccination received 7–89 days before illness onset provided significantly higher protection against hospitalization than original monovalent vaccination only, with an absolute vaccine effectiveness of 52% (39% to 61%) and a relative vaccine effectiveness of 48% (36% to 57%). After 90–179 days, absolute effectiveness waned to 13% (-10% to 31%) and relative effectiveness waned to 17% (-1% to 31%) (figure 2, supplemental figure 2).

**Figure 2.**
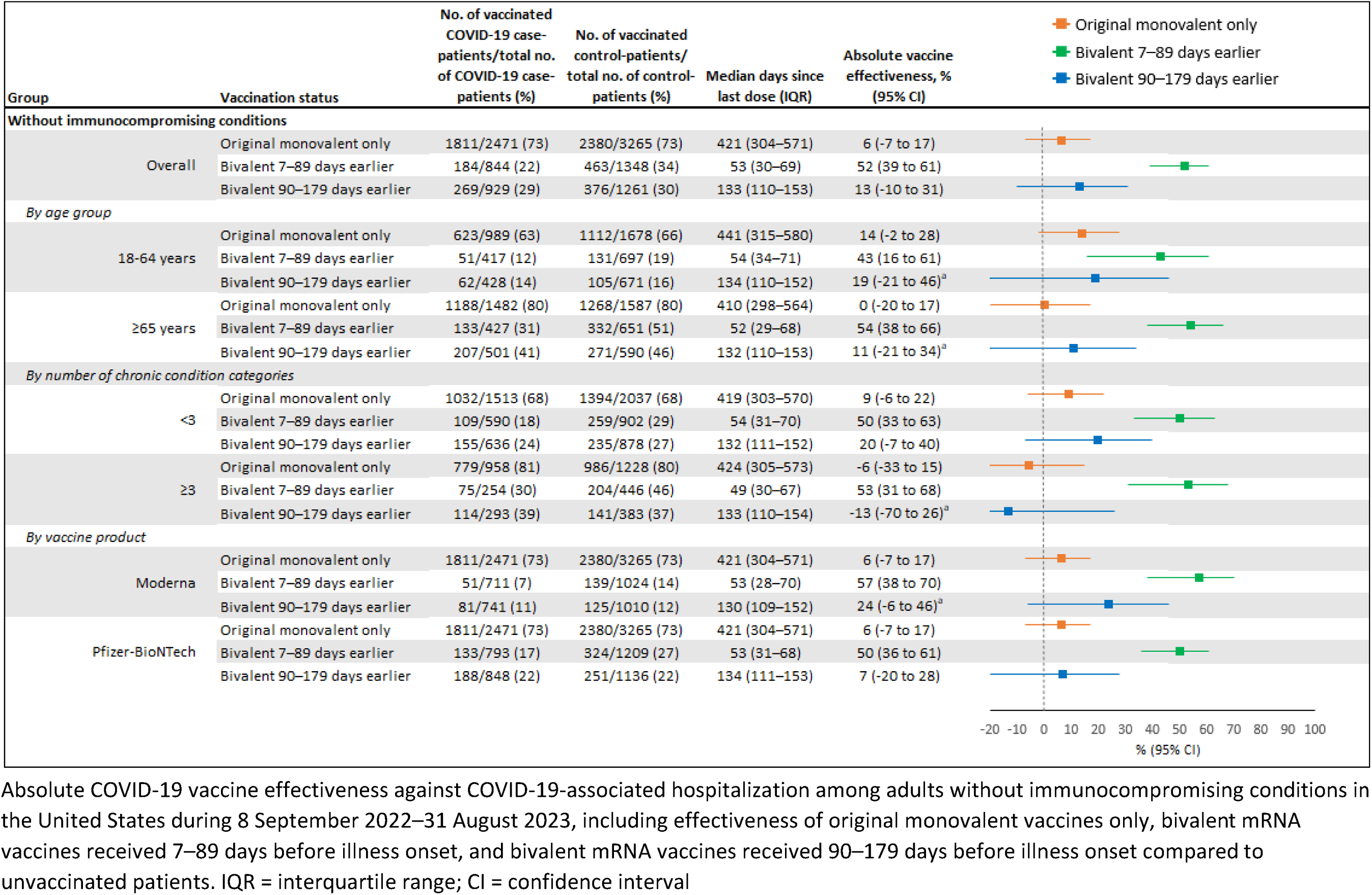
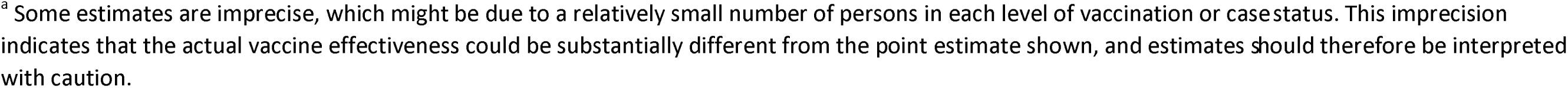

**Figure 3.**
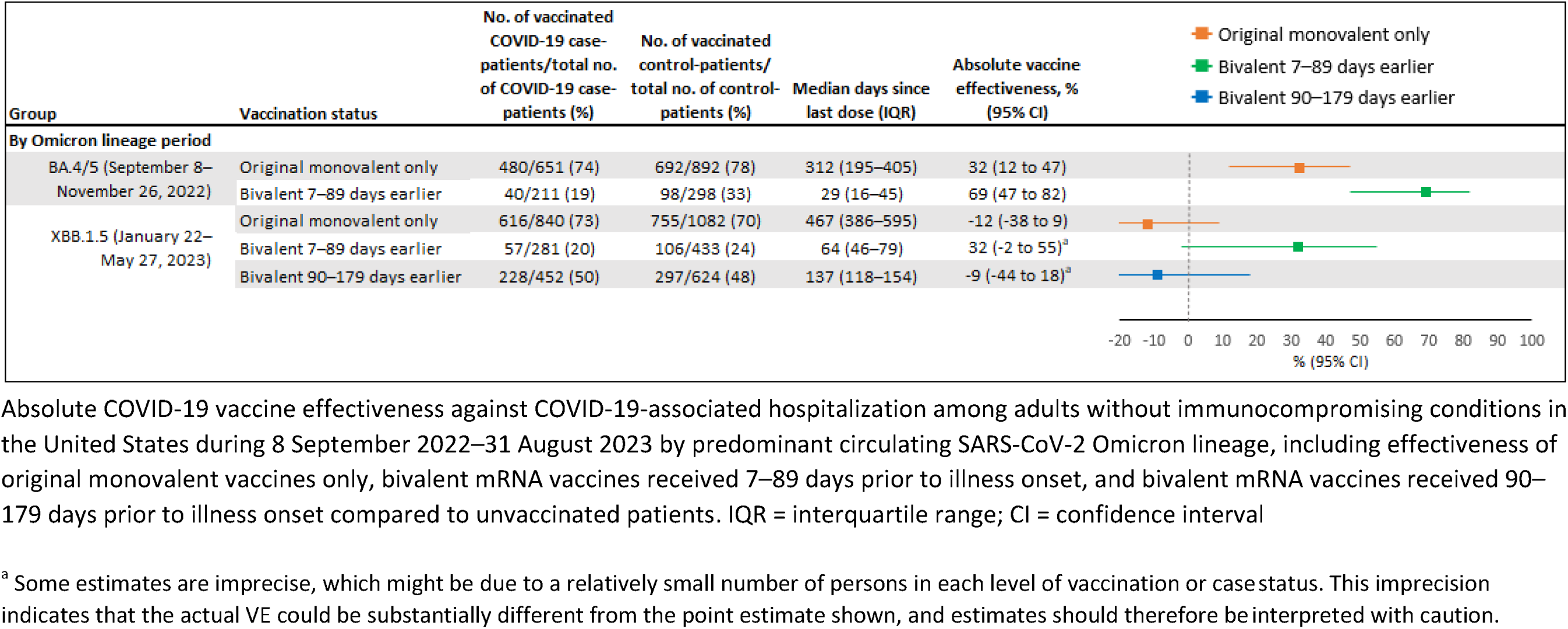

Bivalent vaccination provided increased protection against COVID-19-associated hospitalization 7–89 days after dose receipt compared to original monovalent vaccination only, followed by a decline in point estimates of vaccine effectiveness by 90–179 days across most subgroups, including patients aged ≥65 years, patients with comorbidities in <3 and ≥3 chronic condition categories, Moderna bivalent recipients, and Pfizer bivalent recipients (figure 2, supplemental figure 2). Results were also similar among patients with immunocompromising conditions, although point estimates of bivalent vaccine effectiveness 90–179 days after dose receipt (absolute effectiveness = 45%, 13% to 65%, relative effectiveness = 48%, 28% to 62%) were higher than those among patients without immunocompromising conditions (supplemental figure 5).

Analyses stratified by Omicron lineage period showed a decline in the absolute vaccine effectiveness of original monovalent vaccination only against COVID-19-associated hospitalization from the BA.4/5 period (32%, 12% to 47%) to the XBB.1.5 period (-12%, -38% to 9%). However, the additional protection provided by a bivalent dose received within 7–89 days was similar during both lineage periods. Relative vaccine effectiveness of a bivalent dose received within 7–89 days was 50% (25% to 67%) during the BA.4/5 period and 35% (8% to 54%) during the XBB.1.5 period, although median time since dose receipt among patients who received a bivalent dose within 7–89 days during the BA.4/5 period was 64 days versus 29 days during the XBB.1.5 period. Based on the timing of the lineage periods in relation to the availability of bivalent vaccines, evaluation of bivalent vaccine effectiveness 90–179 days after dose receipt was only possible for the XBB.1.5 period, which showed no remaining protection against COVID-19-associated hospitalization (absolute effectiveness = -9%, -44% to 18%; relative effectiveness = 6%, -16% to 24%).

### Vaccine effectiveness against COVID-19-associated severe in-hospital outcomes

Among 2924 case patients, 632 (21.6%) experienced acute organ failure, 514 (17.8%) were admitted to an ICU, and 281 (9.6%) received IMV or died (table 2). Among 2918 case patients not on home IMV, 1775 (60.8%) received supplemental oxygen therapy and 568 (19.5%) received advanced respiratory support. Compared with case patients who received either original monovalent or bivalent vaccination, unvaccinated case patients were more likely to have advanced respiratory support (p=0.0005), acute organ failure (p=0.0065), ICU admission (p<0.0001), and IMV or death (p<0.0001).

**Table 2.** Severe in-hospital outcomes among adults without immunocompromising conditions admitted to hospital with COVID-19 during 8 September 2022–31 August 2023, by vaccination status. Values are numbers (percentages) of total.

| Clinical outcome | Total | Unvaccinated | Original monovalent only | Bivalent 7–89 days earlier | Bivalent 90–179 days earlier |
| --- | --- | --- | --- | --- | --- |
| Supplemental oxygen therapy<sup>a</sup> | 1775/2918 (60.8) | 406/658 (61.7) | 1086/1807 (60.1) | 120/184 (65.2) | 163/269 (60.6) |
| Advanced respiratory support<sup>b</sup> | 568/2918 (19.5) | 159/658 (24.2) | 323/1807 (17.9) | 34/184 (18.5) | 52/269 (19.3) |
| Acute organ failure<sup>c</sup> | 632/2924 (21.6) | 168/660 (25.5) | 371/1811 (20.8) | 39/184 (21.2) | 54/269 (20.1) |
| ICU admission | 514/2924 (17.8) | 153/660 (23.2) | 281/1811 (15.5) | 38/184 (20.7) | 42/269 (15.6) |
| IMV or death | 281/2924 (9.6) | 92/660 (13.9) | 141/1811 (7.8) | 25/184 (13.4) | 23/269 (8.6) |

Abbreviations: ICU = intensive care unit; IMV = invasive mechanical ventilation
<sup>a</sup> Supplemental oxygen therapy was defined as any receipt of supplemental oxygen for patients with no chronic oxygen needs or escalation of respiratory support for patients on chronic oxygen at baseline. Patients on home IMV were not eligible for this outcome.
<sup>b</sup> Advanced respiratory support was defined as new receipt of high-flow nasal cannula, non-invasive ventilation, or IMV. Patients on home IMV were not eligible for this outcome.
<sup>c</sup> Acute organ failure was defined as respiratory failure (new receipt of high-flow nasal cannula, non-invasive ventilation, or IMV), cardiovascular failure (receipt of vasopressors), or renal failure (new receipt of renal replacement therapy).

The absolute effectiveness of original monovalent vaccination only against COVID-19-associated supplemental oxygen therapy (12%, -2% to 24%) was similar to that against COVID-19-associated hospitalization (6%, -7% to 17%) (figure 4). Point estimates of the absolute effectiveness of original monovalent vaccination only generally increased with outcome severity, up to 51% (34% to 63%) against IMV or death (median time since last dose = 416 days, IQR = 297–564). Compared to the absolute effectiveness of original monovalent doses only, the absolute effectiveness of a bivalent dose received 7–89 days before illness onset was higher against COVID-19-associated supplemental oxygen therapy (original monovalent: 12%, -2% to 24% vs. bivalent: 56%, 42% to 66%), advanced respiratory support (original monovalent: 31%, 15% to 45% vs. bivalent: 66%, 47% to 78%), and acute organ failure (original monovalent: 26%, 8% to 40% vs. bivalent: 61%, 41% to 74%) (figure 4). Estimates of absolute effectiveness for original monovalent vaccination and bivalent vaccination received 7–89 days before illness onset were similar against ICU admission (original monovalent: 36%, 20% to 49% vs. bivalent: 56%, 33% to 71%) and IMV or death (original monovalent: 51%, 34% to 63% vs. bivalent: 61%, 35% to 77%). By 90–179 days after vaccination, point estimates of bivalent vaccine effectiveness against each severe outcome had declined to a level similar to that for original monovalent vaccination only. Trends were similar when assessing relative bivalent effectiveness (supplemental figure 4).

**Figure 4.**
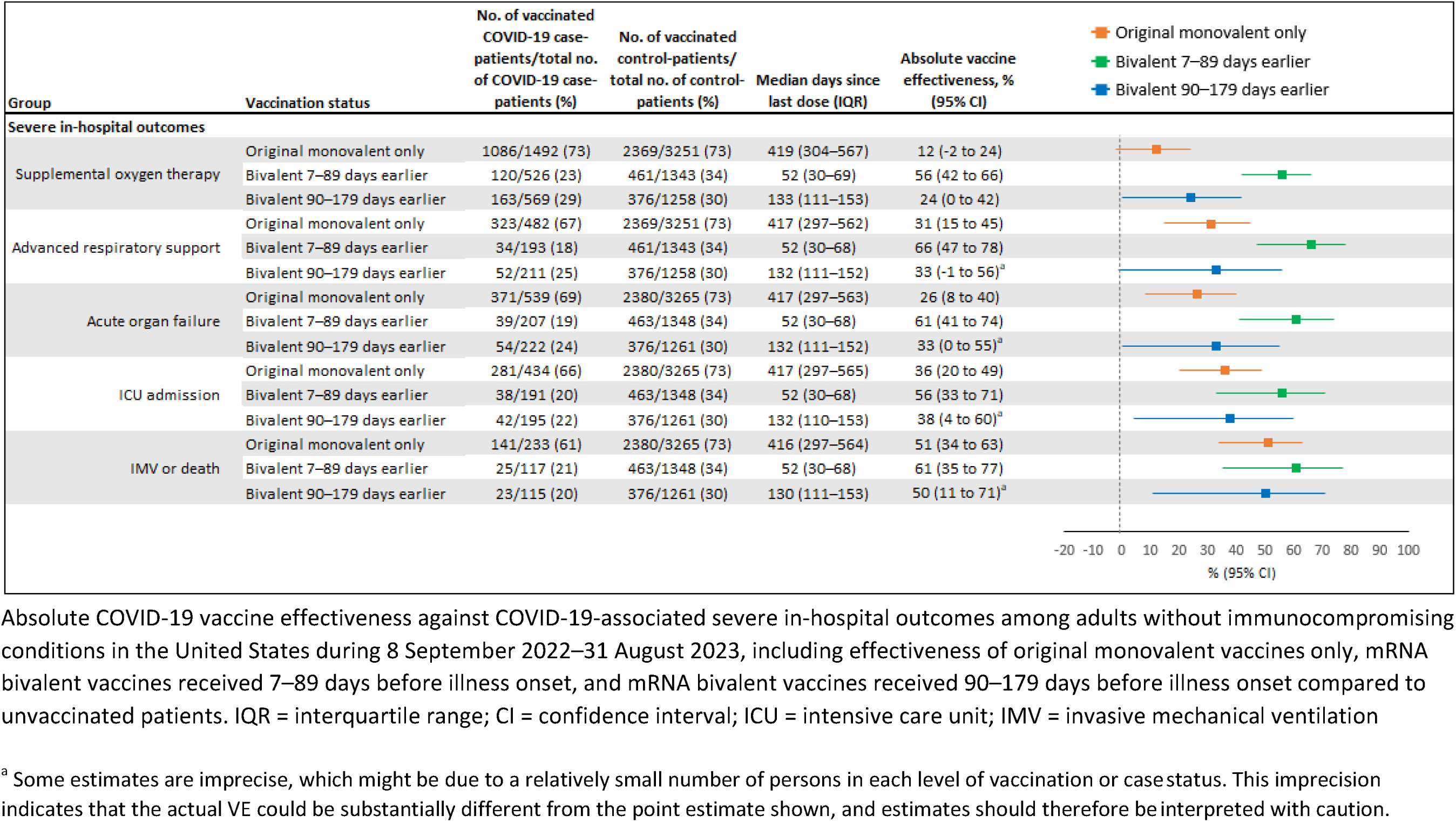

## DISCUSSION

### Principal findings

In this analysis of adults admitted to 26 US hospitals between 8 September 2022 and 31 August 2023, no protection against COVID-19-associated hospitalization remained from original monovalent vaccination only, for which the median time since last dose was >1 year. In contrast, bivalent mRNA COVID-19 vaccination provided protection against COVID-19-associated hospitalization within 3 months of dose receipt, when compared to both unvaccinated patients and patients who received original monovalent doses only. However, protection from bivalent mRNA COVID-19 vaccination waned by 3–6 months after dose receipt. These findings were consistent across key subgroups, including patients aged ≥65 years and patients with multiple comorbidities. Of note, original monovalent vaccination continued to provide durable protection against the most severe in-hospital outcomes, including IMV or death. Bivalent vaccination increased protection against certain severe in-hospital outcomes within 3 months of dose receipt, before declining to a level of protection similar to that remaining from previous original monovalent vaccination. These results support staying up to date with recommended COVID-19 vaccines to optimize protection against both COVID-19-associated hospitalization and severe in-hospital outcomes.

### Comparison with other studies

Our findings are consistent with studies from the US, the United Kingdom, and Finland that have shown waning bivalent COVID-19 vaccine effectiveness against COVID-19-associated hospitalization within 6 months of dose receipt [19–22]. A recent CDC report on US adults without immunocompromising conditions found that the absolute effectiveness of a bivalent vaccine dose against COVID-19-associated hospitalization declined from 62% at 7–59 days after vaccination to 24% after 120–179 days, similar to our results [19]. Declining estimates of bivalent vaccine effectiveness against COVID-19-associated hospitalization may be explained by a number of factors, including the emergence of immune evasive lineages and waning vaccine-induced immunity. Our analysis included both the end of the BA.4/5 predominant lineage period and the XBB.1.5 predominant period in the US. While neutralization studies have identified XBB.1.5 as one of the most immune evasive SARS-CoV-2 lineages to date [12–15], evidence on whether bivalent vaccine effectiveness is lower against XBB.1.5 than against BA.4/5 has been mixed [21,23,24]. We found no significant reduction in the relative effectiveness of a bivalent vaccine dose against COVID-19-associated hospitalization within 7–89 days of receipt during the XBB.1.5 (35%, 8% to 54%) period, compared to the BA.4/5 (50%, 25% to 67%) period. Although the point estimate of effectiveness observed during the XBB.1.5 period was lower than that during the BA.4/5 period, this is likely due, at least in part, to a longer median time since dose receipt among patients who received a bivalent dose within 7–89 days during the XBB.1.5 period (median: 64 days) versus the BA.4/5 period (median: 29 days). Disentangling the potential effects of immune evasion and waning vaccine-induced immunity is challenging, given that both phenomena occur contemporaneously. However, analyses stratified by both lineage period and time since dose receipt showed that during the XBB.1.5 period, point estimates of relative bivalent vaccine effectiveness against hospitalization decreased from 7–89 days (35%, 8% to 54%) after vaccination to 90–179 days (6%, -16% to 24%), although confidence intervals overlapped. Together, these results suggest that waning vaccine-induced immunity may be a stronger driver of declining bivalent vaccine effectiveness than immune evasion from the XBB.1.5 lineage.

Among patients with immunocompromising conditions, bivalent vaccination conferred protection against COVID-19-associated hospitalization within 7–89 days of dose receipt. However, in contrast to the results described for patients without immunocompromising conditions, estimates of bivalent vaccine effectiveness did not show evidence of waning by 90–179 days. Few data are available on bivalent vaccine effectiveness among patients with immunocompromising conditions, but existing studies suggest that COVID-19 vaccine effectiveness wanes in this population as well [19,25,26]. The lack of waning observed in our results may be attributable to behavioral and treatment factors for which we were unable to control in our analysis. Patients with immunocompromising conditions who received a bivalent vaccine dose may have been more likely than unvaccinated patients and patients who received original monovalent vaccination only to practice masking and social distancing for the prevention of SARS-CoV-2 exposure, and to receive outpatient antiviral therapy for the treatment of SARS-CoV-2 infection. The preferential use of these interventions by bivalent recipients could have biased estimates of bivalent vaccine effectiveness away from the null.

Our results also provide important information regarding the effectiveness and durability of COVID-19 vaccination against the most severe in-hospital outcomes. Findings were notable for the long-lasting protection provided by original monovalent COVID-19 vaccination, particularly against ICU admission and IMV or death. In contrast to vaccine-induced immunity against milder infection, which may wane due to decreasing neutralizing antibody levels or immune evasion by emerging variants, immunity against severe outcomes like IMV or death may be mediated by long-lasting memory B- and T-cell responses that are more highly conserved against evolving variants [27–29]. Bivalent vaccination provided additional protection against supplemental oxygen therapy, advanced respiratory support, and acute organ failure within 7–89 days of dose receipt, followed by a decline in protection to a level similar to that remaining from previous original monovalent doses by 90–179 days. However, when compared to original monovalent vaccination only, bivalent vaccination did not significantly increase protection against IMV or death. These results are somewhat in contrast to other studies from Israel, the Nordic countries, and the US demonstrating the effectiveness of bivalent vaccination against death soon after dose receipt [10,11,20,30]. The difference in our results may stem from a combination of high residual protection against IMV or death from original monovalent vaccination and confounding due to prior SARS-CoV-2 infection. Given that prior infection is highly protective against severe outcomes, and may be more likely among patients who are unvaccinated or have a remote history of vaccination compared to recent vaccinees, estimates of vaccine effectiveness against IMV or death may have been biased toward the null [31,32]. Although patient-level data on SARS-CoV-2 infection history were collected for our analysis, these data likely underestimated the prevalence of prior infection in our sample, as many SARS-CoV-2 infections are either undiagnosed or diagnosed through home testing and not reported. Inclusion of these measures in models of bivalent vaccine effectiveness did not appreciably change our results and were not included in the final models.

### Strengths and limitations

This study has several strengths in both its study design and scope. To meet criteria for enrollment, case patients were required to have fever or symptomatic respiratory illness confirmed by either patient interview or electronic health record, limiting the inclusion of case patients who tested positive for SARS-CoV-2, but were hospitalized for reasons other than COVID-19. Ascertainment of vaccination history was also robust, utilizing patient interviews, state registries, and electronic health records to minimize misclassification of vaccination status. Respiratory specimens that tested positive for SARS-CoV-2 underwent viral sequencing, enabling the definition of BA.4/5 and XBB.1.5 lineage periods within our data. An extended study period was included to allow for measurement of bivalent vaccine effectiveness over time up to 6 months after dose receipt. Finally, in addition to presenting estimates of bivalent vaccine effectiveness against COVID-19-associated hospitalization, this analysis also included a range of severe in-hospital outcomes to describe how bivalent vaccines performed against the most severe manifestations of COVID-19.

As with all observational research, this analysis also had limitations. Measures of prior SARS-CoV-2 infection likely underestimated the proportion of patients with prior infection in our sample and were not included in vaccine effectiveness models, as described above. Although vaccine effectiveness estimates were adjusted for patient-level demographic characteristics, calendar time, and geographic region, residual confounding from other factors, including the receipt of COVID-19 antiviral treatments, is also possible. Sample size limitations resulted in wide confidence intervals for some effectiveness estimates and prevented calculation of bivalent vaccine effectiveness against death alone. Lastly, although the systematic collection of data from a large multicenter network increases the external validity of our findings, some results may not be directly generalizable to other settings or populations.

### Policy implications

In light of waning bivalent vaccine effectiveness against COVID-19-associated hospitalization and severe in-hospital outcomes, updated monovalent vaccines against COVID-19 were recommended for US adults in September 2023 [33]. These vaccines were designed against the XBB.1.5 subvariant to more closely match circulating Omicron lineages and to overcome the potential effects of immune imprinting against the ancestral strain of SARS-CoV-2. Receipt of an updated monovalent XBB.1.5 vaccine dose may boost waned vaccine-induced immunity against severe COVID-19 outcomes and improve protection against emerging Omicron lineages. Although morbidity and mortality have decreased considerably since the start of the COVID-19 pandemic, a substantial number of hospitalizations and deaths attributable to COVID-19 continue to occur. During November 2023, approximately 17,000 new COVID-19-associated hospitalizations and 1,300 deaths occurred each week in the US, primarily among older adults [34]. Future monitoring of SARS-CoV-2 epidemiology, the burden of disease in high-risk groups, and the effectiveness of updated monovalent XBB.1.5 vaccines will be critical to informing policy on the need for regular revaccination against COVID-19.

## Conclusions

When compared to original monovalent vaccination only, bivalent mRNA COVID-19 vaccination provided additional protection against COVID-19-associated hospitalization and certain severe in-hospital outcomes within 3 months of dose receipt, after which protection from a bivalent dose declined to a level similar to that remaining from original monovalent vaccination only. Although no residual protection remained from original monovalent vaccination against COVID-19-associated hospitalization, original monovalent vaccination continued to provide durable protection against the most severe COVID-19-associated outcomes >1 year after receipt of the last dose. These results highlight the importance of staying up to date with recommended COVID-19 vaccines to optimize protection against both hospitalization and severe in-hospital outcomes due to COVID-19.

## Data Availability

All data produced in the present study are available upon reasonable request to the authors

## NOTES

### Disclaimer

The findings and conclusions in this report are those of the authors and do not necessarily represent the official position of the Centers for Disease Control and Prevention (CDC).

### Contributions

Guarantors of this work include Dr. Self (protocol and data integrity), Dr. DeCuir (statistical analysis), Dr. Lauring (viral sequencing laboratory methods), and Dr. Chappell (RT-PCR laboratory methods). Contributions of each author include the following. Responsibility for decision to submit the manuscript: DeCuir, Surie, Self. Composed the initial manuscript draft: DeCuir, Surie, Self (the authors only wrote the manuscript without outside assistance). Conceptualization of study methods: DeCuir, Surie, Zhu, Lauring, Martin, Gaglani, McNeal, Ghamande, Peltan, Brown, Ginde, Mohr, Gibbs, Files, Hager, Ali, Prekker, Gong, Mohamed, Johnson, Steingrub, Khan, Busse, Duggal, Wilson, Qadir, Chang, Mallow, Kwon, Exline, Shapiro, Columbus, Vaughn, Ramesh, Safdar, Mosier, Casey, Talbot, Rice, Halasa, Chappell, Grijalva, Baughman, Womack, Rhoads, Swan, Johnson, Lewis, Ellington, McMorrow, Self. Statistical analysis and data management: DeCuir, Zhu, Johnson. Funding acquisition: Self. Critical review of the manuscript for important intellectual content: DeCuir, Surie, Zhu, Lauring, Martin, Gaglani, McNeal, Ghamande, Peltan, Brown, Ginde, Mohr, Gibbs, Files, Hager, Ali, Prekker, Gong, Mohamed, Johnson, Steingrub, Khan, Busse, Duggal, Wilson, Qadir, Chang, Mallow, Kwon, Exline, Shapiro, Columbus, Vaughn, Ramesh, Safdar, Mosier, Casey, Talbot, Rice, Halasa, Chappell, Grijalva, Baughman, Womack, Rhoads, Swan, Johnson, Lewis, Ellington, McMorrow, Self. The corresponding author attests that all listed authors meet authorship criteria and that no others meeting the criteria have been omitted.

### Patient Consent

Not applicable.

### Ethical Approval

This program was approved as a public health surveillance activity with waiver of informed consent by institutional review boards at the US Centers for Disease Control and Prevention (CDC), the program’s coordinating center at Vanderbilt University Medical Center, and each participating site.

### Data Sharing

No additional data are available.

### Transparency

Dr. DeCuir affirms that the manuscript is an honest, accurate, and transparent account of the study being reported; that no important aspects of the study have been omitted; and that any discrepancies from the study as planned have been explained. Dr. Self led the development of the study protocol and participant enrollment. Dr. DeCuir led statistical analysis. Dr. Lauring led viral sequencing work. Dr. Chappell led RT-PCR work. Dr. DeCuir takes responsibility for the work overall.

### Dissemination to participants and related patient and public communities

Results will be disseminated to relevant communities via public health announcements from the US Centers for Disease Control and Prevention, via press releases in the lay press, and public presentations by the investigators.

### Provenance and peer review

Not commissioned; externally peer reviewed.

