## Supplementary Materials for "Durability of protection from original monovalent and bivalent COVID-19 vaccines against COVID-19-associated hospitalization and severe in-hospital outcomes among adults in the United States — September 2022–August 2023"

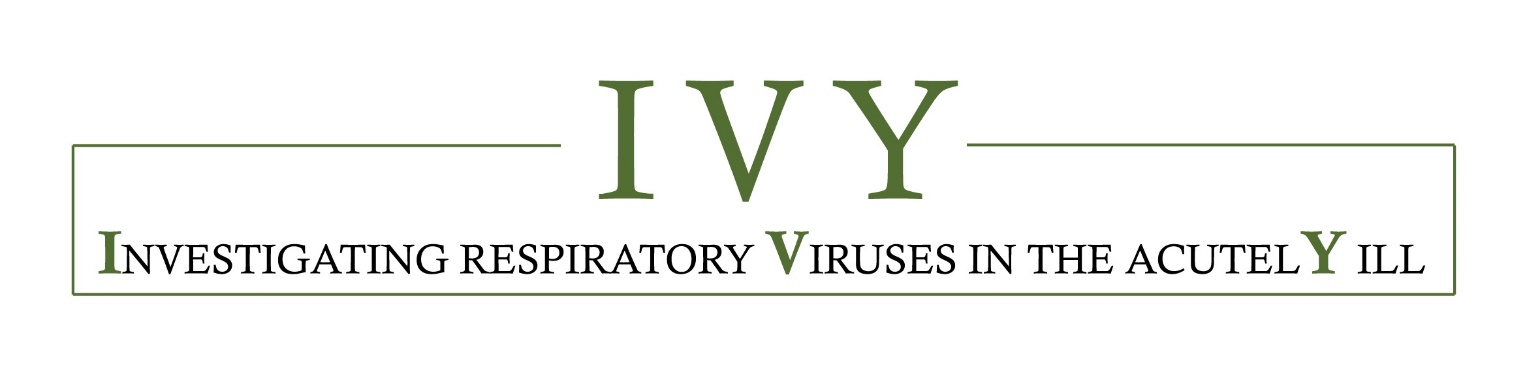

**Supplementary Materials**

Durability of protection from original monovalent and bivalent COVID-19 vaccines against COVID-19-associated hospitalization and severe in-hospital outcomes among adults in the United States — September 2022–August 2023

The IVY Network

**Contents of Supplementary Materials**

**Appendix A**: Investigators and Collaborators ……………………………………………………………………………………………3

**Appendix B**: Supplementary Methods ………………………………………..……………………………………………………………6

1. Chronic medical condition categories obtained through medical record review…….………………6
2. Severe in-hospital outcome definitions…………………………………..……………………………………………..8
3. Laboratory testing methods..…………………………………………………………………………………….…………11

**Supplemental Figure 3.** Relative bivalent vaccine effectiveness against COVID-19-associated hospitalization among adults without immunocompromising conditions in the United States during 8 September 2022–31 August 2023 by predominant circulating SARS-CoV-2 Omicron lineage………………………………………………………………………………………………………………………………………..15

**Supplemental Figure 4.** Relative bivalent vaccine effectiveness against COVID-19-associated severe in-hospital outcomes among adults without immunocompromising conditions in the United States during 8 September 2022–31 August 2023.………………………………………………………….16

**Supplemental Table 1**. Characteristics of adults with immunocompromising conditions by vaccination and COVID-19 status admitted to one of 26 hospitals in 20 US states during 8 September 2022–31 August 2023.………………………………………………………………………………………………17

**Supplemental Figure 5.** COVID-19 vaccine effectiveness against COVID-19-associated hospitalization among adults with immunocompromising conditions in the United States during 8 September 2022–31 August 2023.………………………………………………………………………………………………21

**Supplemental Table 2**. Sensitivity analyses for covariate selection…..………………………………………..22

### Appendix A: Investigators and Collaborators

**Investigators and Collaborators in the Investigating Respiratory Viruses in the Acutely Ill (IVY) Network**

**Baylor, Scott and White, Temple, Texas**

Manjusha Gaglani, Tresa McNeal, Shekhar Ghamande Nicole Calhoun, Kempapura Murthy, Joselyn Cravens, Judy Herrick, Amanda McKillop, Eric Hoffman, Ashley Graves, Martha Zayed, Michael Smith

**Baystate Medical Center, Springfield, Massachusetts**

Jay Steingrub, Lori-Ann Kozikowski, Lesley De Souza, Scott Ouellette

**Beth Israel Medical Center, Boston Massachusetts**

Nathan I. Shapiro, Alessio Barca, Madhavan Das, Jodens Didie, Ana Grafals, Shira Mann, Carlo Ottanelli, Kimberly Redman,

**Centers for Disease Control and Prevention (CDC), Atlanta, Georgia**

Diya Surie, Meredith McMorrow, Jennifer DeCuir, Natalie Thornburg, Brendan Flannery, Nathaniel Lewis, Elizabeth Harker, Katharine Yuengling, Indrani Mukherjee

**Cleveland Clinic, Cleveland, Ohio**

Omar Mehkri, Megan Mitchell, Zachary Griffith, Connery Brennan, Kiran Ashok, Bryan Poynter, Abhijit Duggal

**Emory University, Atlanta, Georgia**

Laurence Busse, Caitlin ten Lohuis, Amy Anderson, Tigist Anemia, Santiago Tovar

**Hennepin County Medical Center, Minneapolis, Minnesota**

Matthew Prekker, Heidi Erickson, Anne Frosch, Audrey Hendrickson, Leyla Taghizadeh, Mary O'Rourke

**Intermountain Medical Center, Murray, Utah**

Ithan Peltan, Samuel Brown, Jenna Lumpkin, Cassie Smith, Shandi Poulsen, Jose Romero

**Johns Hopkins University, Baltimore, Maryland**

David N. Hager, Harith Ali, Richard E. Rothman

**Montefiore Medical Center, Bronx, New York**

Michelle Gong, Amira Mohamed, Rahul Nair, Jen-Ting (Tina) Chen

**Ohio State Medical Center, Columbus, Ohio**

Matthew Exline, Sarah Karow, Maryiam Khan, Madison So, Connor Snyder, Gabrielle Swoope, David Smith, Brooke Lee, Amanie Rasul, Manisha Pathak, Zachery Lewald, Reece Wilson

**Oregon Health and Sciences University, Portland, Oregon**

Akram Khan, Catherine L. Hough, Gopal Allada, Raju Reddy, Shewit P. Giovanni, Bishoy Zakhary, Kinsley Hubel, Sherie Gause, Salil Rajayer

**Stanford University, Stanford, California**

Jennifer G. Wilson, Cynthia Perez, Lily Lau, Ismail Hakki Bekiroglu, Grace Tam, Samantha Ferguson

**University of Arizona**

Jarrod Mosier, Beth Salvagio Campbell, Anitza Lopez, Frances Nagore, Mary Labus

**University of California-Los Angeles, Los Angeles, California**

Nida Qadir, Steven Chang, Cody Tran, Omai Garner, Sukantha Chandrasekaran

**University of Colorado, Aurora, Colorado**

Adit Ginde, Amanda Martinez, Aimee Steinwand, Amy Sullivan, Cori Withers, Jacob Rademacher, Rachel Obradovich

**University of Iowa, Iowa City, Iowa**

Nicholas Mohr, Anne Zepeski, Paul Nassar, Noble Briggs, Jacob Hampton, Cathy Fairfield

**University of Miami, Miami, Florida**

Chris Mallow, Carolina Rivas

**University of Michigan, Ann Arbor, Michigan**

Emily Martin, Arnold Monto, Adam Lauring, EJ McSpadden, Rachel Truscon, Anne Kaniclides, Lara Thomas, Ramsay Bielak, Weronika Damek Valvano, Rebecca Fong, William J. Fitzsimmons, Christopher N. Blair, Julie Gilbert, Leigh Papalambros, Ankur Holz

**University of Washington, Seattle, Washington**

Nicholas Johnson, Vasisht Srinivasan, Christine D. Crider, Kyle A. Steinbock, Thomas C. Paulsen, Layla A. Anderson, Sarah Stucky

**Vanderbilt University Medical Center, Nashville, Tennessee**

Wesley H. Self, H. Keipp Talbot, Carlos Grijalva, Ian Jones, Natasha Halasa, James Chappell, Kelsey Womack, Jillian Rhoads, Adrienne Baughman, Christy Kampe, Jakea Johnson, Sydney Swan, Cassandra Johnson, Yuwei Zhu, Todd Rice, Jonathan Casey, William B. Stubblefield, Yuwei Zhu, Laura L. Short, Lauren J. Ezzell, Margaret E. Whitsett, Rendie E. McHenry, Samarian J. Hargrave, Marcia Blair, Jennifer L. Luther, Claudia Guevara Pulido, Bryan P. M. Peterson, Shanice L. Cummings, Emma Claire Gauthier, Anna C. Jackson, Neekar S. Rashid

**Wake Forest University, Winston-Salem, North Carolina**

D. Clark Files, Kevin Gibbs, Leigha Landreth, Lisa Parks, Fay Nketiaa-Agyepong, Darija Ward, Jacqueline Maycee Cain

**Washington University, St. Louis, Missouri**

Jennie Kwon, Bijal Parikh, David McDonald, Carleigh Samuels, Lucy Vogt, Caroline O’Neil, Alyssa Valencia, Tiffany Hink, Francesca Yerbic, Olivia Arter, Kim Vu

**Baylor, Scott, and White Baylor University Medical Center, Dallas, Texas**

Cristie Columbus, Ashley Bychkowski, Symone Dunkley, Tammy Fisher, Therissa Grefsrud, Mariana Hurutado-Rodriguez, Gabriela Perez

**Henry Ford Health, Detroit, Michigan**

Ivana A Vaughn, Mayur Ramesh, Lois E Lamerato, Kim Beney, Jean Ashley Lava, Melissa Resk, Sindhuja Koneru, Rachna Jayaprakash, Zina Pinderi

**Yale University, New Haven, Connecticut**

Basmah Safdar, Anirudh Goyal, Lauren Delamielleure, Michael Kosover

**Appendix B**: **Supplementary Methods**

1. **Chronic medical condition categories obtained through medical record review**

Condition categories included cardiovascular disease, neurologic disease, pulmonary disease, gastrointestinal disease, endocrine disease, renal disease, hematologic disease, autoimmune disease, and immunocompromising conditions. The main analysis was restricted to patients without immunocompromising conditions and includes estimates of bivalent and original monovalent vaccine effectiveness stratified by number of chronic medical condition categories. A sub-analysis of patients with immunocompromising conditions is included in the supplementary materials.

**Cardiovascular disease**

Heart failure

Peripheral vascular disease that limits mobility

Prior myocardial infarction

Cardiac arrhythmias including atrial fibrillation and ventricular arrhythmias

Valvular heart disease

Hypertension

**Neurologic disease**

Dementia

Prior stroke

Prior transient ischemic attack (TIA, “mini-stroke”)

Brain or spinal cord injury with loss of limb function

Cerebral palsy

Muscular dystrophy

Multiple sclerosis

Myasthenia gravis

Anterolateral sclerosis (ALS)

**Pulmonary disease**

Asthma

Chronic obstructive pulmonary disease

Cystic fibrosis

Pulmonary fibrosis

Pulmonary hypertension

Home oxygen use (except at night for sleep disorder)

Tracheostomy

Home non-invasive ventilation use (except at night for sleep disorder)

Home invasive ventilation use

**Gastrointestinal disease**

Feeding through a tube

Inflammatory bowel disease including Crohn's Disease or Ulcerative Colitis

Cirrhosis (clinical diagnosis of cirrhosis)

Chronic liver disease without cirrhosis

Peptic ulcer disease

**Endocrine disease**

Diabetes mellitus

Adrenal insufficiency

Hypothyroidism

**Kidney disease**

Chronic kidney disease without chronic renal replacement therapy

End stage renal disease on chronic renal replacement therapy (including hemodialysis or peritoneal dialysis)

**Hematologic disease**

Sickle cell disease (all variants)

Coagulopathy or other bleeding disorder, such as hemophilia

Chronic anemia

Thalassemia

**Autoimmune disease**

Systemic Lupus Erythematosus

Rheumatoid arthritis

Psoriasis

Scleroderma

Sarcoidosis

Amyloidosis

Other autoimmune disease

**Immunocompromising conditions**

Active solid organ or hematologic cancer (defined as newly diagnosed cancer or cancer treatment within the past 6 months)

Solid organ transplant

Bone marrow/stem cell transplant

HIV infection

Congenital immunodeficiency syndrome

Use of an immunosuppressive medication within the past 30 days

Splenectomy

Other condition that causes moderate or severe immunosuppression

2. **Severe In-Hospital Outcome Definitions**

The primary outcome in this analysis was COVID-19-associated hospitalization, defined as hospital admission with COVID-19-like illness and laboratory confirmed SARS-CoV-2 infection within 10 days of illness onset and 3 days of hospital admission. The following secondary outcomes were also included to assess vaccine effectiveness against severe COVID-19-associated disease through hospital day 28:

i. COVID-19-associated supplemental oxygen therapy

ii. COVID-19-associated advanced respiratory support

iii. COVID-19-associated acute organ failure

iv. COVID-19-associated intensive care unit (ICU) admission

v. COVID-19-associated invasive mechanical ventilation (IMV) or death

i. COVID-19-associated supplemental oxygen therapy:

Patients who met the definition of COVID-19-associated supplemental oxygen therapy either required supplemental oxygen therapy at any time during the hospitalization through day 28 for those not on chronic oxygen or, for patients on chronic supplemental oxygen (**Table**), required an escalation in respiratory support. Supplemental oxygen therapy could be delivered at any flow rate and by any device; this included standard flow oxygen (flow rate <30 liters/minute), high-flow nasal cannula (HFNC), non-invasive ventilation (NIV), and IMV. Patients on home IMV prior to the acute illness were not eligible for this outcome.

| Classification of in-hospital respiratory outcome based on type of oxygen or respiratory support used chronically (before illness onset) and highest level received through hospital day 28 | | | | | |
| --- | --- | --- | --- | --- | --- |
| Chronic pre-illness oxygen use | Oxygen use during hospital course (highest support) | Is the patient eligible for this outcome? | | | |
|  |  | Supplemental  oxygen therapy | Acute organ failure | Advanced respiratory support | Invasive mechanical ventilation |
| No oxygen use | Standard flow oxygen | Yes | No | No | No |
|  | High-flow nasal cannula (HFNC) | Yes | Yes | Yes | No |
|  | NIV | Yes | Yes | Yes | No |
|  | IMV | Yes | Yes | Yes | Yes |
| Standard flow oxygen | Standard flow oxygen | No | No | No | No |
|  | HFNC | Yes | Yes | Yes | No |
|  | NIV | Yes | Yes | Yes | No |
|  | IMV | Yes | Yes | Yes | Yes |
| Non-invasive mechanical ventilation (NIV) | Standard flow oxygen | No | No | No | No |
|  | HFNC | No | No | No | No |
|  | NIV | No | No | No | No |
|  | IMV | Yes | Yes | Yes | Yes |
| Invasive mechanical ventilation (IMV) | Standard flow oxygen | No | No | No | No |
|  | HFNC | No | No | No | No |
|  | NIV | No | No | No | No |
|  | IMV | No | No | No | No |

ii. COVID-19-associated advanced respiratory support

Patients were classified as having COVID-19-associated advanced respiratory support if they received any of the following during the hospitalization through day 28: HFNC, NIV, or IMV. HFNC was defined as a supplemental oxygen flow rate ≥30 liters per minute. NIV included both continuous positive airway pressure (CPAP) and bilevel positive airway pressure (BiPAP) delivered through a mask. Patients were classified as having NIV use if NIV was received for therapy of the acute illness and not only for treatment of sleep apnea. IMV was defined as positive pressure administered through an endotracheal tube or tracheostomy tube. Patients on home NIV before the acute illness met criteria for the COVID-19-associated advanced respiratory support outcome if they had escalation of respiratory support to IMV in the hospital. Patients on home IMV prior to the acute illness were not eligible for this outcome.

iii. COVID-19-associated acute organ failure

Patients were assessed for receipt of organ support therapies for the respiratory, cardiovascular, and renal systems during the hospitalization through day 28. Patients who were newly treated with any of the following organ support therapies met criteria for the COVID-19-associated acute organ failure outcome:

Organ support for respiratory failure: New receipt of HFNC, NIV, or IMV as described above for COVID-19-associated advanced respiratory support. Patients on home IMV prior to the acute illness could not meet the COVID-19-associated acute organ failure outcome through receipt of organ support for respiratory failure.

Organ support for cardiovascular failure: Intravenous administration of a vasopressor medication by continuous infusion for any duration of time. Vasopressor medications included norepinephrine, vasopressin, epinephrine, dopamine, and phenylephrine.

Organ support for renal failure: New receipt of renal replacement therapy during the hospitalization through hospital day 28. Any type of renal replacement therapy met criteria for this outcome, including hemodialysis and continuous veno-venous hemofiltration. Patients on chronic renal replacement therapy prior to the acute illness could not meet the COVID-19-associated acute organ failure outcome through receipt of organ support for renal failure.

iv. COVID-19-associated intensive care unit (ICU) admission

Patients were classified as having COVID-19-associated ICU admission if they received care in an ICU for any duration of time during the hospitalization through day 28.

v. COVID-19-associated IMV or death

Patients were classified as having COVID-19-associated IMV or death if they received IMV or died during the hospitalization through day 28. IMV was defined as positive pressure administered through an endotracheal tube or tracheostomy tube. Patients on home IMV prior to the acute illness could not meet the COVID-19-associated IMV or death outcome through receipt of in-hospital IMV.

3. **Laboratory Testing Methods**

At the time of participant enrollment, a nasal swab specimen was collected via a fresh swabbing procedure or collection of a residual aliquot in the clinical laboratory. These specimens were frozen at the enrolling site and shipped to Vanderbilt University Medical Center. RT-PCR testing for SARS-CoV-2, influenza, and RSV was completed at Vanderbilt. Specimens with a virus detected were then shipped to the University of Michigan for viral whole genome sequencing. This section describes these laboratory methods.

*SARS-CoV-2 detection by RT-PCR*

For central laboratory pathogen RT-PCR testing at Vanderbilt, total nucleic acid extract from 100 µl of upper respiratory specimen collected in viral transport medium was prepared using the MagNA Pure LC Total Nucleic Acid Isolation Kit (Roche Molecular Systems, Pleasanton, CA) and MagNA Pure 96 automated extraction platform (Roche) or QiaCube HT automated extraction system (Qiagen, Germantown, MD) and QIAamp 96 Virus QiaCube HT kit (Qiagen). Extracts (100 µl eluate volume) were tested by RT-PCR on the StepOnePlus, QuantStudio 3, or QuantStudio 6 Real-Time PCR System (Applied Biosystems, Waltham, MA) for SARS-CoV-2 nucleocapsid (N)-gene N1 and N2 targets and the human RNase P (RNP) gene using the CDC protocol, *CDC 2019-Novel Coronavirus (2019-nCoV) Real-Time RT-PCR Diagnostic Panel* (<https://www.fda.gov/media/134922/download>). RNP served as an endogenous indicator of specimen adequacy and sensor for PCR inhibitors. The pattern of N1, N2, and RNP Ct values served as a basis to assign a qualitative result of *positive*, *not detected*, *inconclusive*, or *invalid specimen* with respect to SARS-CoV-2 RNA according to interpretive criteria delineated in the assay protocol.

*SARS-CoV-2 RNA extraction and whole genome sequencing*

Specimen aliquots with positive RT-qPCR for either the N1 or N2 target with a cycle threshold ≤40 at Vanderbilt University Medical Center laboratory were shipped to the University of Michigan on dry ice. RNA was extracted from 200µl transport media with the Thermo Fisher MagMAX Viral Pathogen II Isolation Kit on a KingFisher instrument and eluted in a 50µl volume. Extracted RNA was reverse transcribed with Lunascript RT Supermix (NEB). For each sample, 2 µl of master-mix was added to 8µl of RNA template and incubated at 25°C for 2 min, 55°C for 20 min, 95C for 1 min. Viral cDNA was amplified in two multiplex PCR reactions with the ARTIC Network (currently version 5.3.2) primer pools and protocol using the Q5 Hot Start High-Fidelity DNA Polymerase Master-mix (NEB) with the following thermocycler protocol: 98˚C for 30 s, then 35 cycles of 95˚C for 15 s, 63˚C for 5 min. Reaction products for a given sample were pooled together in equal volumes. Sequencing libraries were prepared by ligation of the appropriate Illumina or Oxford Nanopore Technologies (ONT) adaptor sets. Three negative control wells (1 HeLa RNA, 2 water) were included on each 96 well RNA harvest plate and carried through the entire process. Barcoded libraries were pooled and sequenced in batches of 96 (GridION instrument) or 384 (Illumina Nextseq 1000, 2x300 P1 flow cell). A run was repeated from RNA harvest on if any of the negative controls have >30x read coverage over 10% of the genome. PANGO lineage was assigned on genomes with >80% coverage using Pangolin v2.4.1 (https://pangolin.cog-uk.io, citation in main text). Genomes with >90% coverage were uploaded to GISAID (<https://www.gisaid.org/>).

*Influenza detection by RT-PCR*

For central laboratory pathogen RT-PCR testing at Vanderbilt, total nucleic acid extract from 100 µl of upper respiratory specimen collected in viral transport medium was prepared using the MagNA Pure LC Total Nucleic Acid Isolation Kit (Roche Molecular Systems, Pleasanton, CA) and MagNA Pure 96 automated extraction platform (Roche) or QiaCube HT automated extraction system (Qiagen, Germantown, MD) and QIAamp 96 Virus QiaCube HT kit (Qiagen). Extracts (100 µl eluate volume) were tested by RT-PCR on the StepOnePlus, QuantStudio 3, or QuantStudio 6 Real-Time PCR System (Applied Biosystems, Waltham, MA) for influenza A and B using the CDC Human Influenza Virus Real-Time RT-PCR Diagnostic Panel, Influenza A/B Typing Kit (VER 2). Each specimen also was tested for RNase P (RNP) as a marker of specimen adequacy and sensor for PCR inhibitors using TaqPath 1-Step RT-qPCR Master Mix, CG (Applied Biosystems). PCR reactions consisted of 45 amplification cycles, and Ct values of any magnitude were deemed positive when represented by a characteristic specific amplification curve. Absence of influenza A and/or B detection in specimens registering RNP Ct values ≥38 was considered inconclusive for the undetected virus(es).

*RSV detection by RT-PCR*

For central laboratory pathogen RT-PCR testing at Vanderbilt, total nucleic acid extract from 100 µl of upper respiratory specimen collected in viral transport medium was prepared using the MagNA Pure LC Total Nucleic Acid Isolation Kit (Roche Molecular Systems, Pleasanton, CA) and MagNA Pure 96 automated extraction platform (Roche) or QiaCube HT automated extraction system (Qiagen, Germantown, MD) and QIAamp 96 Virus QiaCube HT kit (Qiagen). Extracts (100 µl eluate volume) were tested by RT-PCR on the StepOnePlus, QuantStudio 3, or QuantStudio 6 Real-Time PCR System (Applied Biosystems, Waltham, MA) for a pan-RSV matrix gene target using Superscript III Platinum One-Step Quantitative RT-PCR System with ROX passive reference dye (Invitrogen, Waltham, MA) and a screening set of primers and probes (forward: GGCAAATATGGAAACATACGTGAA; reverse: TCTTTTTCTAGGACATTGTAYTGAACAG; probe: FAM-CTGTGTATGTGGAGCCTTCGTGAAGCT-BHQ-1) (Biosearch Technologies, Petaluma, CA). Each specimen also was tested for RNase P (RNP) as a marker of specimen adequacy and sensor for PCR inhibitors using TaqPath 1-Step RT-qPCR Master Mix, CG (Applied Biosystems). PCR reactions consisted of 45 amplification cycles, and Ct values of any magnitude were deemed positive when represented by a characteristic specific amplification curve. Absence of RSV detection in specimens registering RNP Ct values ≥40 was considered inconclusive for viral RNA.

**Appendix C: Supplementary Figures and Tables**

**Supplemental Figure 1.** Participant flow diagram

**11797** patients admitted during

8 September 2022–31 August 2023

**2374** Excluded^a^:

• **61** Did not meet eligibility criteria

• **261** SARS-CoV-2 testing indeterminate or not done

• **20** Case patient received a positive influenza test result

• **20** Control patient received a positive influenza test result

• **297** Influenza testing indeterminate or not done

• **45** Case patient received a positive RSV test result

• **432** Unknown vaccination status

• **19** Immunocompetent and received 5 monovalent doses

• **412** Received one monovalent Moderna, Pfizer, or Novavax dose

• **81** Received last dose <7 days before illness onset

• **800** Received bivalent dose ≥180 days before illness onset

• **18** Received >1 bivalent dose

• **49** Immunocompetent, age <50, and received >3 monovalent doses

• **14** Withdrew

**9423** patients

**2395** patients with immunocompromising conditions included in sub-analysis (supplement)

**7028** patients without immunocompromising conditions

included in main analysis

**1451** Control patients:

• **213** Unvaccinated

• **915** Monovalent only

• **165** Bivalent 7–89 days earlier

• **158** Bivalent 90–179 days earlier

**944** Case patients:

• **157** Unvaccinated

• **645** Monovalent only

• **64** Bivalent 7–89 days earlier

• **78** Bivalent 90–179 days earlier

**4104** Control patients:

• **885** Unvaccinated

• **2380** Monovalent only

• **463** Bivalent 7–89 days earlier

• **376** Bivalent 90–179 days earlier

**2924** Case patients:

• **660** Unvaccinated

• **1811** Monovalent only

• **184** Bivalent 7–-89 days earlier

• **269** Bivalent 90–179 days earlier

**^a^** Exclusions not mutually exclusive

**Supplemental Figure 2.**

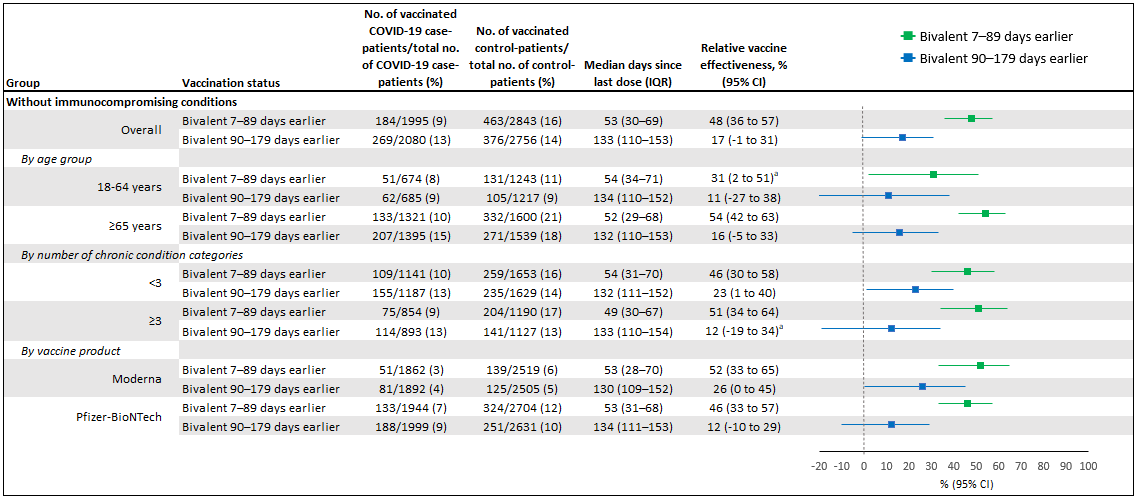

Relative bivalent vaccine effectiveness against COVID-19-associated hospitalization among adults without immunocompromising conditions in the United States during 8 September 2022–31 August 2023, including effectiveness of bivalent mRNA vaccines received 7–89 days before illness onset and bivalent mRNA vaccines received 90–179 days before illness onset compared to patients who received original monovalent vaccines only. IQR = interquartile range; CI = confidence interval

^a^ Some estimates are imprecise, which might be due to a relatively small number of persons in each level of vaccination or case status. This imprecision indicates that the actual VE could be substantially different from the point estimate shown, and estimates should therefore be interpreted with caution.

**Supplemental Figure 3.**

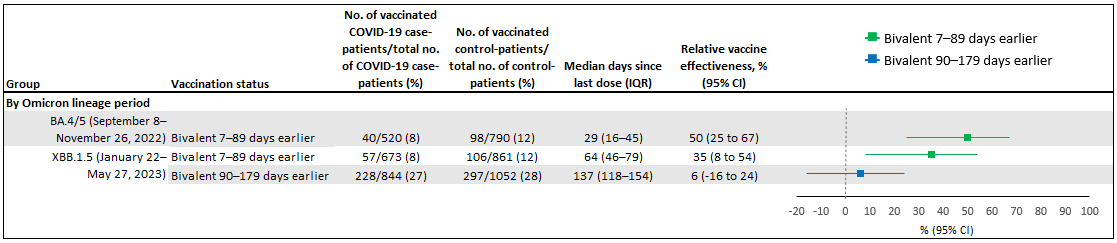

Relative bivalent vaccine effectiveness against COVID-19-associated hospitalization among adults without immunocompromising conditions in the United States during 8 September 2022–31 August 2023 by predominant circulating SARS-CoV-2 Omicron lineage, including effectiveness of bivalent mRNA vaccines received 7–89 days prior to illness onset and bivalent mRNA vaccines received 90–179 days prior to illness onset compared to patients who received original monovalent vaccines only. IQR = interquartile range; CI = confidence interval

**Supplemental Figure 4.**

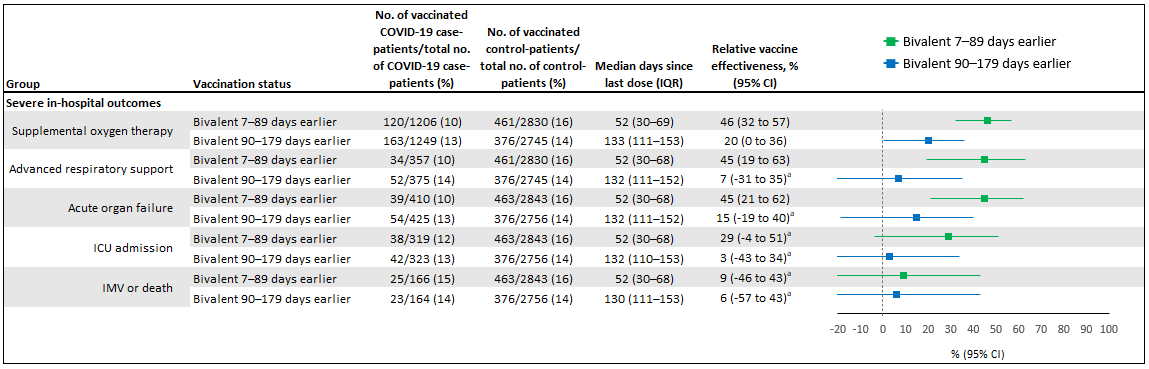

Relative bivalent vaccine effectiveness against COVID-19-associated severe in-hospital outcomes among adults without immunocompromising conditions in the United States during 8 September 2022–31 August 2023, including effectiveness of bivalent mRNA vaccines received 7–89 days before illness onset and bivalent mRNA vaccines received 90–179 days before illness onset compared to patients who received original monovalent vaccines only. IQR = interquartile range; CI = confidence interval; ICU = intensive care unit; IMV = invasive mechanical ventilation

^a^ Some estimates are imprecise, which might be due to a relatively small number of persons in each level of vaccination or case status. This imprecision indicates that the actual VE could be substantially different from the point estimate shown, and estimates should therefore be interpreted with caution.

**Supplemental Table 1.** Characteristics of adults with immunocompromising conditions by vaccination and COVID-19 status admitted to one of 26 hospitals in 20 US states during 8 September 2022–31 August 2023. Values are numbers (column percentages) unless stated otherwise

| **Characteristics** | **Total**  **(n=2395)** | **Vaccination status** | | | |  | **COVID-19 status** | |
| --- | --- | --- | --- | --- | --- | --- | --- | --- |
|  |  | **Unvaccinated**  **(n=370)** | **Monovalent only**  **(n=1560)** | **Bivalent 7**–**89 days earlier**  **(n=229)** | **Bivalent 90**–**179 days earlier**  **(n=236)** |  | **COVID-19 case patients**  **(n=944)** | **Control patients**  **(n=1451)** |
| Median (IQR) age (years) | 63 (52–72) | 55 (41–67) | 64 (53–72) | 66 (56–74) | 67 (57–75) |  | 64 (53–73) | 63 (51–71) |
| Age group (years) |  |  |  |  |  |  |  |  |
| 18–49 | 517/2395 (21.6) | 137/370 (37.0) | 309/1560 (19.8) | 35/229  (15.3) | 36/236  (15.3) |  | 191/944 (20.2) | 326/1451 (22.5) |
| 50–64 | 761/2395 (31.8) | 118/370 (31.9) | 510/1560 (32.7) | 67/229  (29.3) | 66/236  (28.0) |  | 289/944 (30.6) | 472/1451 (32.5) |
| ≥65 | 1117/2395 (46.6) | 115/370  (31.1) | 741/1560 (47.5) | 127/229 (55.5) | 134/236 (56.8) |  | 464/944 (49.2) | 653/1451 (45.0) |
| Female | 1153/2395 (48.1) | 188/370 (50.8) | 765/1560 (49.0) | 98/229  (42.8) | 102/236  (43.2) |  | 463/944 (49.1) | 690/1451 (47.6) |
| Race and ethnicity |  |  |  |  |  |  |  |  |
| White, non-Hispanic | 1429/2395 (59.7) | 204/370 (55.1) | 912/1560 (58.5) | 153/229 (66.8) | 160/236 (67.8) |  | 550/944 (58.3) | 879/1451 (60.6) |
| Black or African American, non-Hispanic | 485/2395 (20.3) | 89/370  (24.1) | 310/1560 (19.9) | 46/229  (20.1) | 40/236  (17.0) |  | 195/944 (20.7) | 290/1451 (20.0) |
| Hispanic or Latino, any race | 301/2395 (12.6) | 52/370  (14.1) | 213/1560 (13.7) | 14/229  (6.1) | 22/236  (9.3) |  | 129/944 (13.7) | 172/1451 (11.9) |
| Other race, non-Hispanic^a^ | 95/2395 (4.0) | 9/370  (2.4) | 67/1560  (4.3) | 9/229  (3.9) | 10/236  (4.2) |  | 40/944  (4.2) | 55/1451 (3.8) |
| Other^b^ | 85/2395 (3.6) | 16/370  (4.3) | 58/1560  (3.7) | 7/229  (3.1) | 4/236  (1.7) |  | 30/944  (3.2) | 55/1451 (3.8) |
| HHS region^c^ |  |  |  |  |  |  |  |  |
| 1 | 318/2395 (13.3) | 28/370  (7.6) | 219/1560 (14.0) | 34/229  (14.9) | 37/236  (15.7) |  | 139/944 (14.7) | 179/1451 (12.3) |
| 2 | 85/2395 (3.6) | 17/370  (4.6) | 60/1560  (3.9) | 4/229  (1.8) | 4/236  (1.7) |  | 34/944  (3.6) | 51/1451 (3.5) |
| 3 | 139/2395 (5.8) | 17/370  (4.6) | 82/1560  (5.3) | 15/229  (6.6) | 25/236  (10.6) |  | 78/944  (8.3) | 61/1451 (4.2) |
| 4 | 313/2395 (13.1) | 62/370  (16.8) | 200/1560 (12.8) | 33/229  (14.4) | 18/236  (7.6) |  | 134/944 (14.2) | 179/1451 (12.3) |
| 5 | 569/2395 (23.8) | 85/370  (23.0) | 363/1560 (23.3) | 60/229  (26.2) | 61/236  (25.9) |  | 198/944 (21.0) | 371/1451 (25.6) |
| 6 | 265/2395 (11.1) | 48/370  (13.0) | 176/1560 (11.3) | 21/229  (9.2) | 20/236  (8.5) |  | 97/944 (10.3) | 168/1451 (11.6) |
| 7 | 217/2395 (9.1) | 48/370  (13.0) | 138/1560 (8.9) | 20/229  (8.7) | 11/236  (4.7) |  | 65/944  (6.9) | 152/1451 (10.5) |
| 8 | 158/2395 (6.6) | 28/370  (7.6) | 95/1560  (6.1) | 16/229  (7.0) | 19/236  (8.1) |  | 54/944  (5.7) | 104/1451 (7.2) |
| 9 | 261/2395 (10.9) | 27/370  (7.3) | 190/1560 (12.2) | 18/229  (7.9) | 26/236  (11.0) |  | 115/944 (12.2) | 146/1451 (10.1) |
| 10 | 70/2395 (2.9) | 10/370  (2.7) | 37/1560  (2.4) | 8/229  (3.5) | 15/236  (6.4) |  | 30/944  (3.2) | 40/1451 (2.8) |
| No. of organ systems with chronic conditions , median (IQR)^d^ | 2 (1–3) | 2 (1–3) | 2 (1–3) | 2 (1–4) | 3 (1–4) |  | 2 (1–3) | 2 (1–3) |
| Immunocompromising conditions |  |  |  |  |  |  |  |  |
| Active solid organ cancer^e^ | 813/2342 (34.7) | 134/361 (37.1) | 545/1530 (35.6) | 61/219  (27.9) | 73/232  (31.5) |  | 280/916 (30.6) | 533/1426 (37.4) |
| Active hematologic cancer^e^ | 397/2342 (17.0) | 69/361  (19.1) | 267/1530 (17.5) | 30/219  (13.7) | 31/232  (13.4) |  | 161/916 (17.6) | 236/1426 (16.6) |
| Solid organ transplant | 565/2342 (24.1) | 57/361  (15.8) | 364/1530 (23.8) | 75/219  (34.3) | 69/232  (29.7) |  | 269/916 (29.4) | 296/1426 (20.8) |
| Bone marrow/stem cell transplant | 122/2342 (5.2) | 24/361  (6.7) | 80/1530  (5.2) | 9/219  (4.1) | 9/232  (3.9) |  | 42/916  (4.6) | 80/1426 (5.6) |
| HIV infection | 191/2341 (8.2) | 42/360  (11.7) | 106/1530  (6.9) | 19/219  (8.7) | 24/232  (10.3) |  | 70/916  (7.6) | 121/1425 (8.5) |
| Congenital immunodeficiency syndrome | 4/2341 (0.2) | 0/360  (0.0) | 3/1530  (0.2) | 1/219  (0.5) | 0/232  (0.0) |  | 3/916  (0.3) | 1/1425  (0.1) |
| Use of Immunosuppressive medication in the past 30 days | 1644/2394 (68.7) | 245/369 (66.4) | 1068/1560 (68.5) | 161/229 (70.3) | 170/236 (72.0) |  | 648/944 (68.6) | 996/1450 (68.7) |
| Splenectomy | 32/2341 (1.4) | 7/360  (1.9) | 20/1530  (1.3) | 2/219  (0.9) | 3/232  (1.3) |  | 19/916  (2.1) | 13/1425  (1.0) |
| Other immunocompromising condition | 119/2341 (5.1) | 17/360  (4.7) | 78/1530  (5.1) | 13/219  (5.9) | 11/232  (4.7) |  | 45/916  (4.9) | 74/1425 (5.2) |
| Previous SARS-CoV-2 infection^f^ |  |  |  |  |  |  |  |  |
| Any previous SARS-CoV-2 infection | 731/2395 (30.5) | 122/370 (33.0) | 469/1560 (30.1) | 65/229  (28.4) | 75/236  (31.8) |  | 243/944 (25.7) | 488/1451 (33.6) |
| Previous Omicron variant infection | 490/2395 (20.5) | 79/370  (21.4) | 314/1560 (20.1) | 43/229  (18.8) | 54/236  (22.9) |  | 163/944 (17.3) | 327/1451 (22.5) |
| COVID-19 status |  |  |  |  |  |  |  |  |
| Case patients | 944/2395 (39.4) | 157/370 (42.4) | 645/1560 (41.4) | 64/229  (28.0) | 78/236  (33.1) |  | 944/944 (100.0) | -- |
| Control patients | 1451/2395 (60.6) | 213/370 (57.6) | 915/1560 (58.7) | 165/229 (72.1) | 158/236 (67.0) |  | -- | 1451/1451 (100.0) |
| Vaccination status |  |  |  |  |  |  |  |  |
| Unvaccinated | 370/2395 (15.5) | 370/370 (100.0) | -- | -- | -- |  | 157/944 (16.6) | 213/1451 (14.7) |
| Monovalent only | 1560/2395 (65.1) | -- | 1560/1560 (100.0) | -- | -- |  | 645/944 (68.3) | 915/1451 (63.1) |
| Bivalent 7–89 days earlier | 229/2395  (9.6) | -- | -- | 229/229 (100.0) | -- |  | 64/944  (6.8) | 165/1451 (11.4) |
| Bivalent 90–179 days earlier | 236/2395  (9.9) | -- | -- | -- | 236/236 (100.0) |  | 78/944  (8.3) | 158/1451 (10.9) |
| Bivalent vaccine product received^g^ |  |  |  |  |  |  |  |  |
| Moderna | 137/465 (29.5) | -- | -- | 63/229  (27.5) | 74/236  (31.4) |  | 37/142 (26.1) | 100/323 (31.0) |
| Pfizer | 328/465 (70.5) | -- | -- | 166/229 (72.5) | 162/236 (68.6) |  | 105/142 (73.9) | 223/323 (69.0) |

Abbreviations: IQR = interquartile range; HHS = U.S. Department of Health and Human Services

^a^ “Other race, non-Hispanic” includes Asian, Native American or Alaska Native, and Native Hawaiian or other Pacific Islander; these groups were combined due to small counts.

^b^ “Other” includes patients who self-reported their race and ethnicity as “Other” and those for whom race and ethnicity were unknown.

^c^ Hospitals by HHS region included *Region 1*: Baystate Medical Center (Springfield, Massachusetts), Beth Israel Deaconess Medical Center (Boston, Massachusetts), and Yale University (New Haven, Connecticut); *Region 2*: Montefiore Medical Center (New York, New York); *Region 3*: Johns Hopkins Hospital (Baltimore, Maryland); *Region 4*: Emory University Medical Center (Atlanta, Georgia), University of Miami Medical Center (Miami, Florida), Vanderbilt University Medical Center (Nashville, Tennessee), and Wake Forest University Baptist Medical Center (Winston-Salem, North Carolina); *Region 5*: Cleveland Clinic (Cleveland, Ohio), Hennepin County Medical Center (Minneapolis, Minnesota), Henry Ford Health (Detroit, Michigan), The Ohio State University Wexner Medical Center (Columbus, Ohio), and University of Michigan Hospital (Ann Arbor, Michigan); *Region 6*: Baylor Scott & White Medical Center (Temple, Texas) and Baylor University Medical Center (Dallas, Texas); *Region 7*: Barnes-Jewish Hospital (St. Louis, Missouri) and University of Iowa Hospitals (Iowa City, Iowa); *Region 8*: Intermountain Medical Center (Murray, Utah), UCHealth University of Colorado Hospital (Aurora, Colorado), and University of Utah (Salt Lake City, Utah); *Region 9*: Stanford University Medical Center (Stanford, California), Ronald Reagan UCLA Medical Center (Los Angeles, California), and University of Arizona Medical Center (Tucson, Arizona); and *Region 10*: Oregon Health & Science University Hospital (Portland, Oregon) and University of Washington (Seattle, Washington).

^d^ Chronic medical condition categories include autoimmune, cardiovascular, endocrine, gastrointestinal, hematologic, neurologic, pulmonary, and renal diseases.

^e^ Active cancer was defined as newly diagnosed cancer or cancer treatment in the past 6 months.

^f^ Previous SARS-CoV-2 infection was defined as any self-reported or documented SARS-CoV-2 infection that occurred before the episode of illness for which the patient was enrolled into the IVY Network. Previous Omicron infection was defined as any self-reported or documented SARS-CoV-2 infection that occurred from December 26, 2021 until the episode of illness for which the patient was enrolled into the IVY Network.

### ^g^ Proportion denominators restricted to bivalent recipients only.

**Supplemental Figure 5.**

**A)**

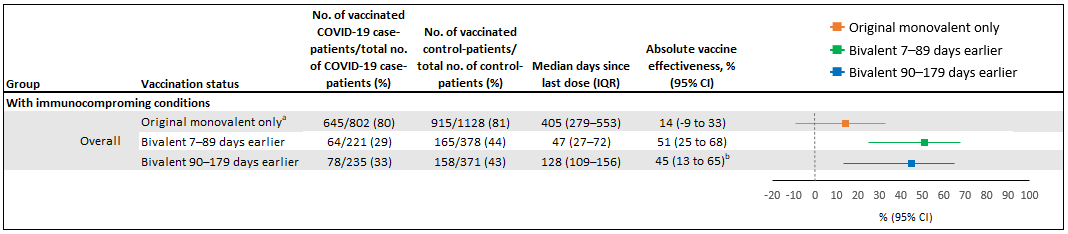

**B)**

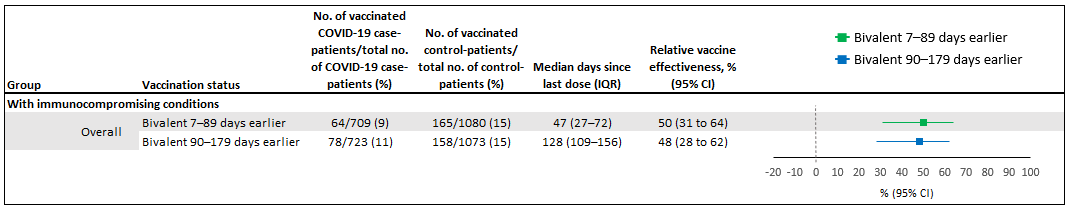

COVID-19 vaccine effectiveness against COVID-19-associated hospitalization among adults with immunocompromising conditions in the United States during 8 September 2022–31 August 2023, including A) estimates of absolute vaccine effectiveness compared to unvaccinated patients and B) estimates of relative vaccine effectiveness compared to patients who received original monovalent vaccines only. IQR = interquartile range; CI = confidence interval

^a^ Patients with immunocompromising conditions were classified as vaccinated with original monovalent doses only if they received any combination of any combination of 1–5 doses of original monovalent vaccine: mRNA-1273 [Moderna], BNT162b2 [Pfizer-BioNTech], Ad26.COV2.S [Janssen], NVX-CoV2373 [Novavax]).

^b^ Some estimates are imprecise, which might be due to a relatively small number of persons in each level of vaccination or case status. This imprecision indicates that the actual VE could be substantially different from the point estimate shown, and estimates should therefore be interpreted with caution.

**Supplemental Table 2.** Sensitivity analyses for covariate selection

| **Models** | **Absolute vaccine effectiveness^a^, %** | | |
| --- | --- | --- | --- |
|  | **Original monovalent only** | **Bivalent 7**–**89 days earlier** | **Bivalent 90**–**179 days earlier** |
| Primary (prespecified) model^b^ | 6 | 52 | 13 |
| Primary model + number of chronic medical condition categories^c^ | 6 | 50 | 12 |
| Primary model + any previous SARS-CoV-2 infection^d^ | 5 | 52 | 13 |
| Primary model + previous Omicron infection^e^ | 4 | 52 | 13 |

Absolute vaccine effectiveness against COVID-19-associated hospitalization among adults without immunocompromising conditions in the United States during 8 September 2022–31 August 2023 was estimated in a series of models to evaluate additional variables as potential covariates, beyond those that were included in the prespecified model. None of these other variables resulted in an absolute change in the adjusted odds ratio of >5% when added to the prespecified model and were not included in the final models.

^a^ Absolute vaccine effectiveness was estimated using unvaccinated patients used as the reference group.

^b^ The primary model was adjusted for age (18-49, 50-64, ≥65), sex (male, female), self-reported race and Hispanic ethnicity (non-Hispanic white, non-Hispanic Black, Hispanic or Latino, non-Hispanic other race, other), admission date in biweekly intervals, and U.S. Department of Health and Human Services region of the admitting hospital.

^c^ Chronic medical condition categories include autoimmune, cardiovascular, endocrine, gastrointestinal, hematologic, neurologic, pulmonary, and renal diseases.

^d^ Previous SARS-CoV-2 infection was defined as any self-reported or documented SARS-CoV-2 infection that occurred before the episode of illness for which the patient was enrolled into the IVY Network.

^e^ Previous Omicron infection was defined as any self-reported or documented SARS-CoV-2 infection that occurred from December 26, 2021 until the episode of illness for which the patient was enrolled into the IVY Network.
